# Global genomics in over 4 million individuals prioritizes therapeutic targets for heart failure and its subtypes

**DOI:** 10.64898/2026.08.13.26360411

**Authors:** Danielle Rasooly, Gina M. Peloso, Claudia Giambartolomei, Hannah L. Nicholls, Chang Liu, Nay Aung, Hesam Dashti, Kai Gravel-Pucillo, Jaime Berumen, Jesús Alegre-Díaz, Pablo Kuri-Morales, Roberto Tapia-Conyer, VA Million Veteran Program, John Whittaker, Peter W. F. Wilson, Lawrence S. Phillips, Kelly Cho, J. Michael Gaziano, Yan V. Sun, Jason M. Torres, Alexandre C. Pereira, Juan P. Casas, Jacob Joseph

## Abstract

Heart failure (HF) is a leading cause of morbidity and mortality. We conducted multi-ancestry genome-wide association studies of 345,687 HF cases (4,468,166 individuals), and 47,192 and 46,934 cases of HF with preserved (HFpEF) and reduced ejection fraction (HFrEF), respectively, integrating plasma proteomics and multi-tissue transcriptomics to identify druggable targets. Across HF, HFrEF, and HFpEF, we identified 383 loci (166 novel) and 568 genes (375 novel). Eleven novel genes are targets of approved or investigational cardiovascular therapies, supporting indication expansion of aldosterone synthase inhibitors (*CYP11B2*) and type-II activin receptor antagonists (*ACVR2A*) to HF. Six cardiomyopathy genes were novel for HF and associated with cardiac structure and function. We identified nearly 100 genes involved in food intake and energy expenditure; metabolism of fatty acids, glucose, and branched-chain amino acids; and mitochondrial proteome, sustaining myocardial energy production. Our findings highlight the primordial role of metabolic pathways and adipokines as therapeutic targets for HF management.

## Introduction

Heart failure (HF) remains a major contributor to global morbidity and mortality, and its growing burden underscores the need to identify novel therapeutic targets. Despite recent improvements in the management of HF with reduced ejection fraction (HFrEF)^1,2^, substantial unmet therapeutic needs persist. These needs are even more pronounced in HF with preserved ejection fraction (HFpEF), which has fewer established disease-modifying therapies. This therapeutic gap is further compounded by the global rising prevalence of obesity, a strong risk factor to both HFrEF and HFpEF.

Since the first large-scale genetic study of HF six years ago^3^, there has been a rapid expansion in the identification of putative drug targets; however, most studies have been conducted in predominantly European-descent populations^3–6^, and few have explicitly distinguished between HFrEF and HFpEF^5,7^. The burden of HF is unevenly distributed across populations^8,9^, with underrepresented racial and ethnic groups experiencing higher incidence, earlier onset, and worse outcomes. These populations remain understudied in genetic research, limiting the discovery of ancestry-shared mechanisms and constraining opportunities for therapeutic target identification.

To address these gaps, we performed large-scale multi-ancestry genome-wide association studies (GWAS) and plasma proteome-and multi-tissue transcriptome-wide Mendelian randomization (MR) studies of HF, HFpEF, and HFrEF, comprising 345,687 individuals with HF (47,192 with HFpEF and 46,934 with HFrEF) and >4.12 million control individuals, spanning European, African, Asian, and Admixed American ancestries. To improve the generalizability of HF genetic discoveries, we incorporated substantial representation of non-European ancestry populations, including 55,854 individuals with HF (16,023 with HFpEF and 14,160 with HFrEF) and 970,872 control individuals of African, Asian, and Admixed American ancestries. To strengthen the validity of our findings, we triangulated orthogonal evidence spanning cardiac imaging, pathway analysis, RNA expression profiling, functional annotation, and therapeutic tractability assessment.

## RESULTS

### Genomic loci associated with HF, HFrEF, and HFpEF

We meta-analyzed the Million Veteran Program (MVP), HERMES Consortium, Biobank Japan, the Global Biobank Meta-analysis Initiative, the All of Us Research Program, the Mexico City Prospective Study, and FinnGen R12, comprising 4,468,166 individuals, including 345,687 HF cases. The HFpEF analysis included 47,192 cases and a total of 1,514,078 individuals; the HFrEF analysis included 46,934 cases and a total of 1,695,762 individuals **(Figure 1A)**. Study characteristics for all included cohorts are provided in **Table S1**.

**Figure 1.**
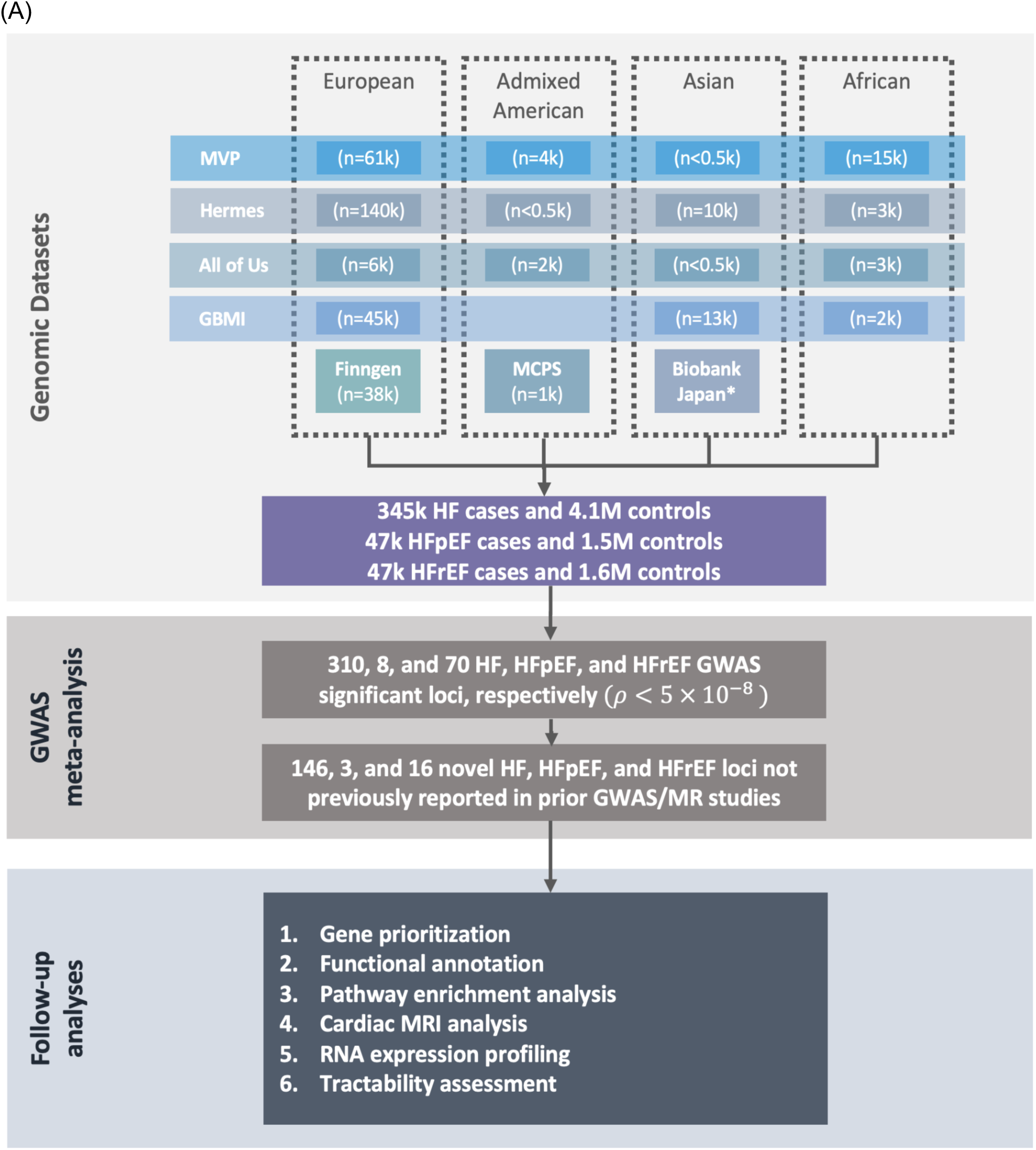
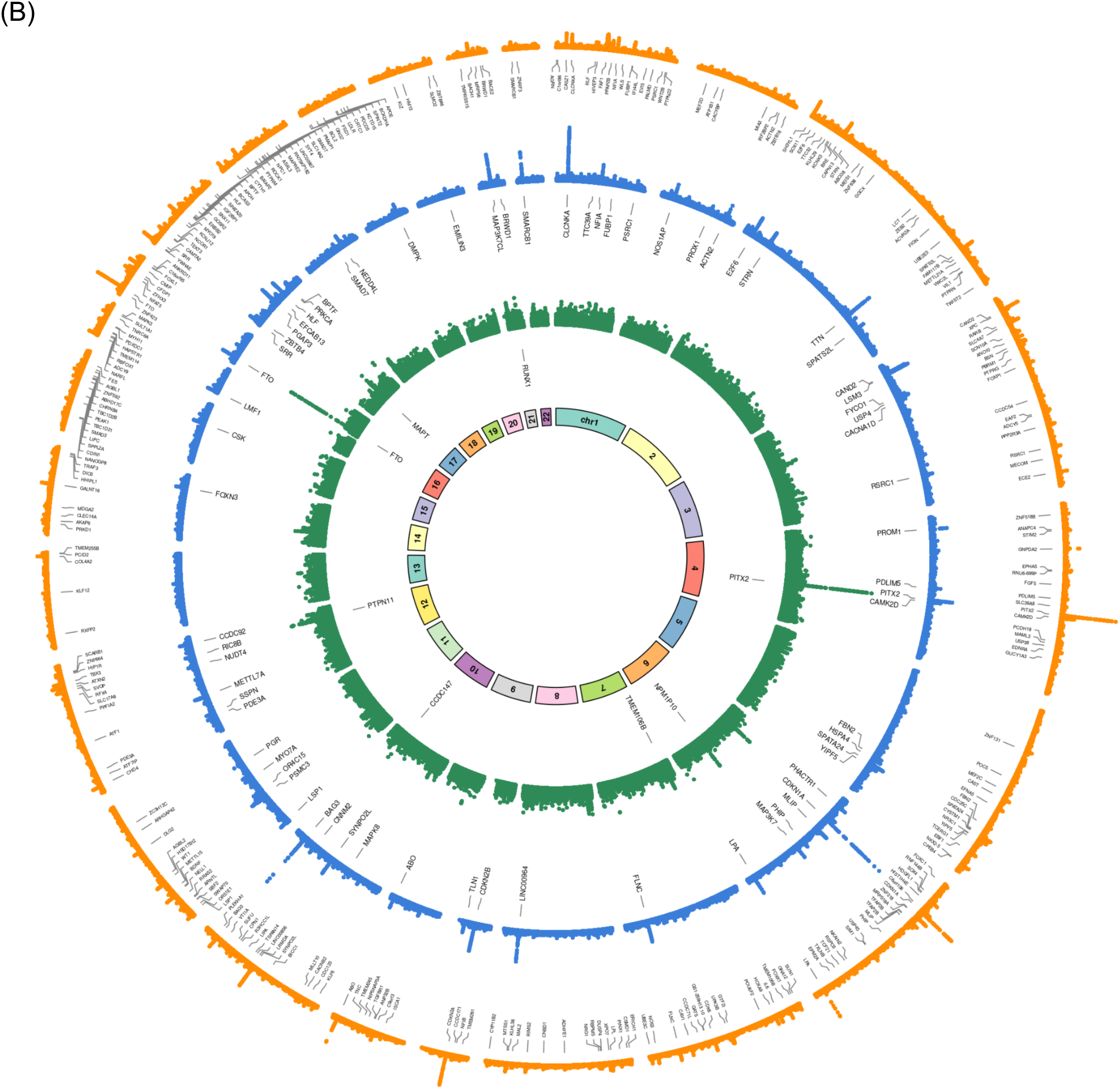
Study overview and GWAS results of the multi-ancestry analysis of HF, HFrEF, and HFpEF. (A) We performed a multi-ancestry genome-wide association study (GWAS) of heart failure (HF) including 345,687 cases and 4,122,479 controls (total *n* = 4,468,166) from the Million Veteran Program, HERMES Consortium, Global Biobank Meta-analysis Initiative, All of Us Research Program, Mexico City Prospective Study and FinnGen Release 12. Subtype-specific analyses were conducted for HF with preserved ejection fraction (HFpEF; 47,192 cases and 1,466,886 controls; total *n* = 1,514,078) and HF with reduced ejection fraction (HFrEF; 46,934 cases and 1,648,828 controls; total *n* = 1,695,762). For the HFrEF and HFpEF GWAS analyses, genomic data from Biobank Japan (BBJ) were included. This is indicated by an asterisk because BBJ was excluded from the overall HF analysis, having already been represented in the HERMES study. Across analyses, individuals of European, Asian, African and Admixed American descent were included. Genome-wide significance was defined as *P* < 5 × 10⁻⁸. Novel loci were determined through comparison with previously reported HF GWAS and MR studies. Follow-up analyses were conducted on genome-wide significant signals to further characterize their functional and biological relevance. (B) Circular Manhattan plot of multi-ancestry HF, HFrEF, and HFpEF GWAS results. Circular Manhattan plot showing results for multi-ancestry all-cause HF (orange), HFrEF (blue) and HFpEF (green). The innermost ring denotes chromosome numbers. Loci are labeled by the gene most frequently implicated across four gene-mapping approaches (cS2G, V2G, nearest gene and MAGMA); in cases of ties, a single representative gene is displayed for visualization purposes.

We identified 415 independent variants (p < 5 × 10⁻⁸) across 310 genomic loci for HF, defined by merging LD blocks within 250 kb (**Table S2**). Manhattan and quantile-quantile plots are shown in **Figure 1B and Figure S1**. Of these, 146 loci (47.1%) represent novel associations not previously reported in GWAS or MR studies of HF, HFpEF, and HFrEF **(Figure S2)**. For HFpEF, eight loci (3 novel) reached genome-wide significance; novel genes included *RUNX1*, *NPM1P10*, and *CCDC147* **(Table S3)**. For HFrEF, 70 loci (16 novel) reached genome-wide significance (**Table S4**).

### Gene mapping, fine-mapping and functional analyses

We mapped GWAS loci to candidate genes using cs2g, variant-to-gene (V2G), MAGMA, and nearest gene approaches, prioritizing the gene most frequently assigned across methods and retaining multiple genes when assignment remained unclear. 72% of loci showed concordance across multiple methods and 37% showed concordance across all methods (**Table S2-S4**). Ensembl VEP annotation showed that most GWAS variants were noncoding and were classified as modifier impact, while the coding variants were largely predicted to be tolerated or benign (**Table S5**). We performed fine-mapping using 90% credible sets across all HF, HFrEF, and HFpEF loci, resolving 46 loci to a single candidate variant and an additional 100 loci to ≤5 candidate variants; the remaining loci retained broader credible sets (>5 variants) (**Table S6**). Exploratory bioinformatic functional enrichment analysis showed that HFrEF was enriched for sarcomere and contractile processes and HFpEF for inflammatory pathways (**Table S7**). Among GWAS-identified genes, 11 were highly expressed (top 25%) exclusively in cardiac or vascular tissue, while 21 were highly expressed exclusively in skeletal muscle, kidney, or adipose tissue, and 133 genes were highly expressed in at least one of these tissues (**Figure S3-S4**).

### MR-QTLs for HF, HFpEF, and HFrEF

We performed proteome-and transcriptome two-sample MR analyses (**Figure S5**) using plasma protein abundance and gene expression across 49 tissues for 17,079 protein-coding genes. We identified 521 genes whose genetically predicted expression or circulating protein level was associated with HF, HFrEF, and HFpEF (referred to thereafter as “MR genes”) that met the Bonferroni significance threshold (p < 1.6 × 10⁻⁶) **(Table S8-S11)**. A total of 467 genes were identified for HF, 12 for HFpEF, and 130 for HFrEF (**Table S8**). Of 521 unique MR genes, 119 (22.8%) were also the assigned GWAS gene. **Figure S6**-**S7** shows the distribution across HF phenotypes. For HFpEF, *HLA-DRB1*, *SPPL2C, and FSTL4* were novel, while *MYOZ1*, *HLA-DQA1*, *TMEM106B* was previously reported as HF genes and here refined to HFpEF (**Table S9**). For HFrEF, 53 genes were novel, and for 11 genes, we refined a previously reported HF association to HFrEF (**Table S10**). Among MR genes, 14 were highly expressed (top 25%) exclusively in cardiac or vascular tissue, while 48 were highly expressed exclusively in skeletal muscle, kidney, or adipose tissue, and 233 genes were highly expressed in at least one of these tissues (**Table S12** and **Figure S3-S4**).

The MR instruments had a median (IQR) F-statistic of 80.0 (49.1, 165.6), indicating strong instrument strength. For associations with ≥3 instruments (25 HF, 1 HFpEF, 5 HFrEF), no evidence of directional pleiotropy was detected using MR-Egger intercept tests (**Table S8-10**). A total of 215 MR genes out of 521 were in LD (r² ≥ 0.4) with previously reported variants from HF, HFrEF, or HFpEF GWAS or MR studies (**Table S13** and **S14**).

### Orthogonal evidence supporting genes identified by GWAS and MR

We aggregated evidence from multiple sources into a composite score: animal genetics, OMIM/ClinGen, cardiac MRI traits relevant to HF, prior GWAS and MR evidence, and OMIM/ClinGen and animal genetics evidence on genes directly linked to our gene findings (**Table S15 to S18**). We identified 272 of 732 (37.2%) genes with at least one supporting line of orthogonal evidence (**Figure S8**). The top 5% of genes by score included established CM genes such as *BAG3*, *TTN*, *FLNC*, and *ACTN2*, supporting the scoring framework. *FKBP7*, a novel HFrEF gene identified by MR (β = −0.23; P = 1.63 × 10⁻⁹), achieved the highest score, with supporting evidence from animal genetics exhibiting HF phenotypes and associations with >15 cardiac MRI measures of LV contractility, remodeling, RV remodeling, and diastolic dysfunction.

### Genetically supported drug targets

Using GWAS and MR, we identified 125 HF, HFpEF, and HFrEF genes annotated as druggable or potentially druggable by Open Targets (**Table S19**). Seventeen genes (11 novel and 6 known) are targets of approved or investigational therapies for cardiovascular disorders (**Table 1**). Novel genes include *CYP11B2*, the target of aldosterone synthase inhibitors, approved for resistant hypertension and under evaluation for incident HF in the Prevent-HF Study^10^, and *ACVR2A*, the target of type-II activin receptor antagonists approved for pulmonary hypertension and currently evaluated for obesity and combined post-and precapillary pulmonary hypertension associated with HFpEF (CpcPH-HFpEF)^11^. We rediscovered the targets of established drugs used in HF management: *GUCY1A1* (vericiguat), *ATP1B2* (digoxin), and *PDE3A* (milrinone) for severe HF. We identified *PPP1CB*, which encodes the PP1 beta catalytic subunit (PP1β), one of the core catalytic isoforms of protein phosphatase 1 (PP1), which is the target of AB-1002, an investigational gene therapy candidate expressing constitutively active, truncated form of protein phosphatase inhibitor-1 to inhibit PP1, currently being tested in a Phase 2 trial^12^. At FDR of 5%, we identified the missense variant rs1800437/*GIPR* associated with HF (p=2×10⁻⁶, β=-0.02). Glucose-dependent insulinotropic polypeptide receptor (*GIPR*) is one of the targets of tirzepatide, which showed beneficial effects in individuals with obese HFpEF^13^. Additional gene targets of licensed or investigational cardiovascular drugs are described in **Table 1**.

**Table 1.** Genetically supported heart failure (HF) and HF subtypes drug targets with approved and investigational therapies.

| Gene | HF Phenotype | Genetic evidence for HF | Novelty | Therapeutic modality | Drug Name | Condition | FDA-approved |
| --- | --- | --- | --- | --- | --- | --- | --- |
| **Cardiovascular/Neurohormonal†** |  |  |  |  |  |  |  |
| *CYP11B2* | HF | GWAS | Novel | Small molecule | Aldosterone synthase inhibitors (Baxdrostat; Lorundrostat) | Hypertension | Yes (Baxdrostat); Yes (Lorundrostat) |
| *GUCY1A1* | HF | GWAS, MR | Novel | Small molecule | Vericiguat | HFrEF | Yes |
| *ATP1B2* | HF | MR | Novel | Small molecule | Digoxin, Istaroxime | Atrial fibrillation, acute HF | Yes (Digoxin), No (Istaroxime) |
| *PDE3A* | HFrEF, HF | GWAS | Novel | Small molecule | Milrinone | HF, pulmonary hypertension | Yes |
| *NRG1* | HF | GWAS, MR | Novel | Antibody | JK07 | HF | No |
| *ENPEP* | HF | MR | Novel | Small molecule | QGC-001 (Firibastat) | Hypertension, HF | No |
| *ADM* | HF | MR | Known | Antibody | Enibarcimab | Septic shock, HF | No |
| *EDNRA* | HF | GWAS | Known | Small molecule | Tezosentan, Bosentan | Pulmonary hypertension, diastolic HF | Yes (Bosentan); No (Tezosentan) |
| *CACNB2* | HF | GWAS, MR | Known | Small molecule | Calcium channel blockers, Dronedarone | Hypertension, coronary artery disease, atrial fibrillation, congestive HF | Yes |
| *RXFP2* | HF | GWAS | Known | Protein | Serelaxin** | Coronary artery disease, cardiovascular disease, HF, chronic kidney disease | No |
| **Obesity/Metabolism\*** |  |  |  |  |  |  |  |
| *LEPR* | HF | MR | Novel | Protein | Metreleptin | Lipodystrophy, obesity | Yes |
| *ACVR2A* | HF | GWAS, MR | Novel | Protein | Sotatercept, Bimagrumab | Pulmonary arterial hypertension, obesity | Yes (Sotatercept); No (Bimagrumab) |
| *MC4R* | HF | GWAS | Known | Protein | Setmelanotide | Severe obesity | Yes |
| **Lipids** |  |  |  |  |  |  |  |
| *PCSK9* | HF | MR | Novel | Antibody, siRNA | Evolocumab, Alirocumab, Inclisiran | Hyperlipidemia | Yes |
| *LPA* | HF, HFrEF | GWAS, MR | Known | ASO, siRNA | Pelacarsen, Olpasiran, Zerlasiran, Lepodisiran | ASCVD | No |
| **Inflammation** |  |  |  |  |  |  |  |
| *IL6\** | HF | GWAS | Novel | Antibody | Ziltiviekimab, Pacibekitug | ASCVD, HFpEF | No |
| **Other Mechanisms** |  |  |  |  |  |  |  |
| *PPP1CB* | HF | MR | Novel | AAV | AB-1002 | HF | No |

\*We identified rs1800437/*GIPR* as associated with HF at FDR of 5% ( $p=2E-6$ ). This is one of the targets of tirzepatide that showed beneficial effects in individuals with obese HFpEF<sup>13</sup>. †We identified *AGT* as associated with HF at an FDR of 5% in MR analyses. *AGT* is a target of zilebesiran. \*\*Serelaxin is an agonist of the *RXFP1* receptor and a weak agonist of the *RXFP2* receptor. The natural ligand for *RXFP2* with the highest affinity is *INSL3*.

### Shared genetic architecture with dilated and hypertrophic cardiomyopathy

We integrated previously reported cardiomyopathy (CM) genes from GWAS and comprehensive gene panels with our HF genetic analyses, identifying 14 novel and 26 known cardiomyopathy (CM) genes associated with HF, HFrEF, or HFpEF (**Figure S9**). Notable novel genes included *SSPN*, *ALPK3*, and *HSPA4* (**Figure 2A**). *ALPK3*, an essential component of the sarcomeric M-band, was associated with HF and measures of LV remodeling, including LVM, LVEDV, and LVESV (**Figure 2B**). *SSPN*, a HFrEF gene, stabilizes the dystrophin-glycoprotein complex (DGC), which preserves sarcolemmal integrity in cardiomyocytes; *DAG1*, another DGC component, was identified as a HF gene (**Figure 2C**). *SSPN* was associated with measures of cardiac contractility (LV-GCS and LV-GRS) and LV remodeling (LVESV), supporting its association with HFrEF. HSPA4, which encodes a protective cardiomyocyte chaperone, was associated with both HFrEF and HF. It showed associations with LV contractility (LVEF and LV-GCS), LV remodeling (LVM), and RV remodeling (RVESV), supporting its link to HFrEF and suggesting relevance to diastolic dysfunction.

**Figure 2.**
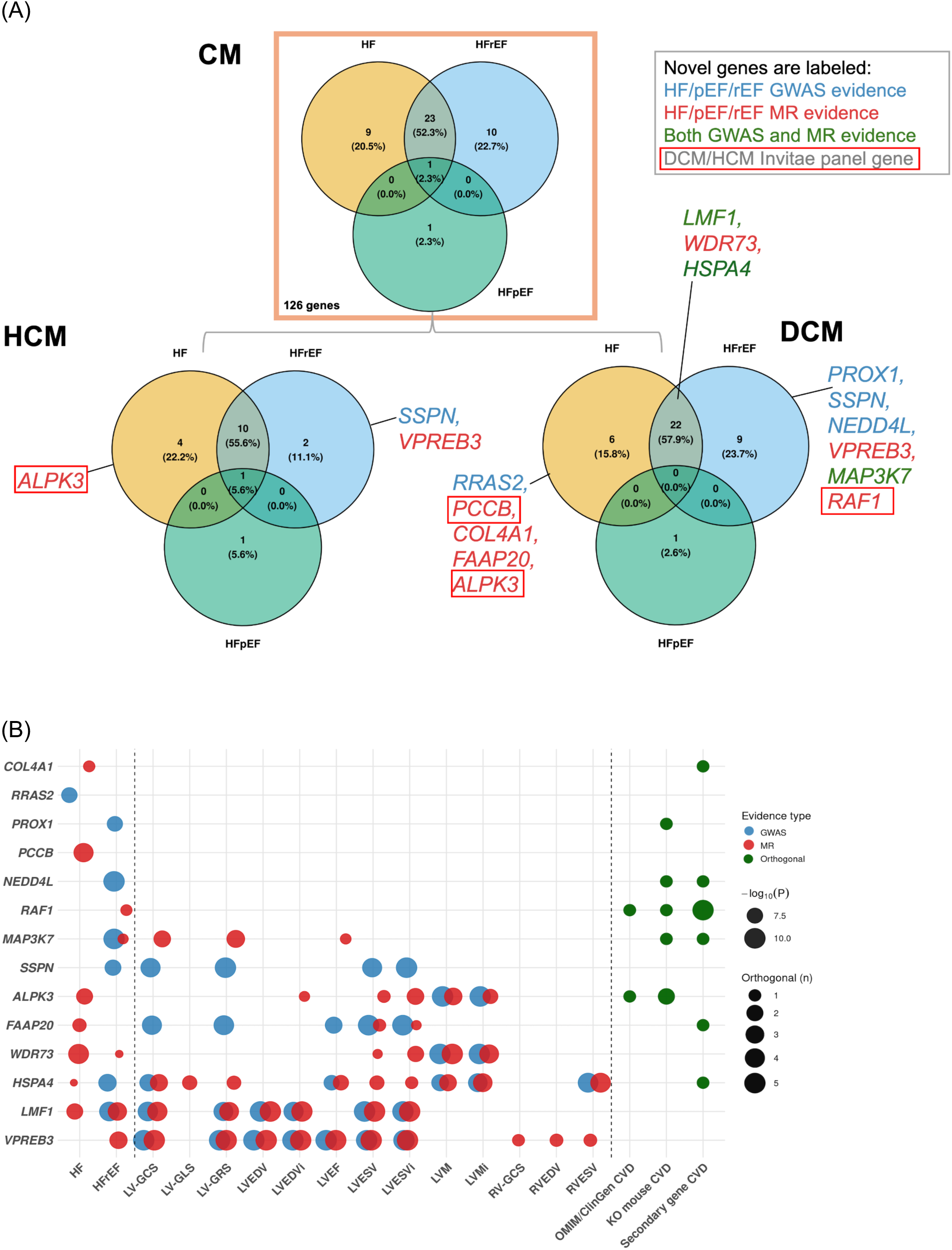
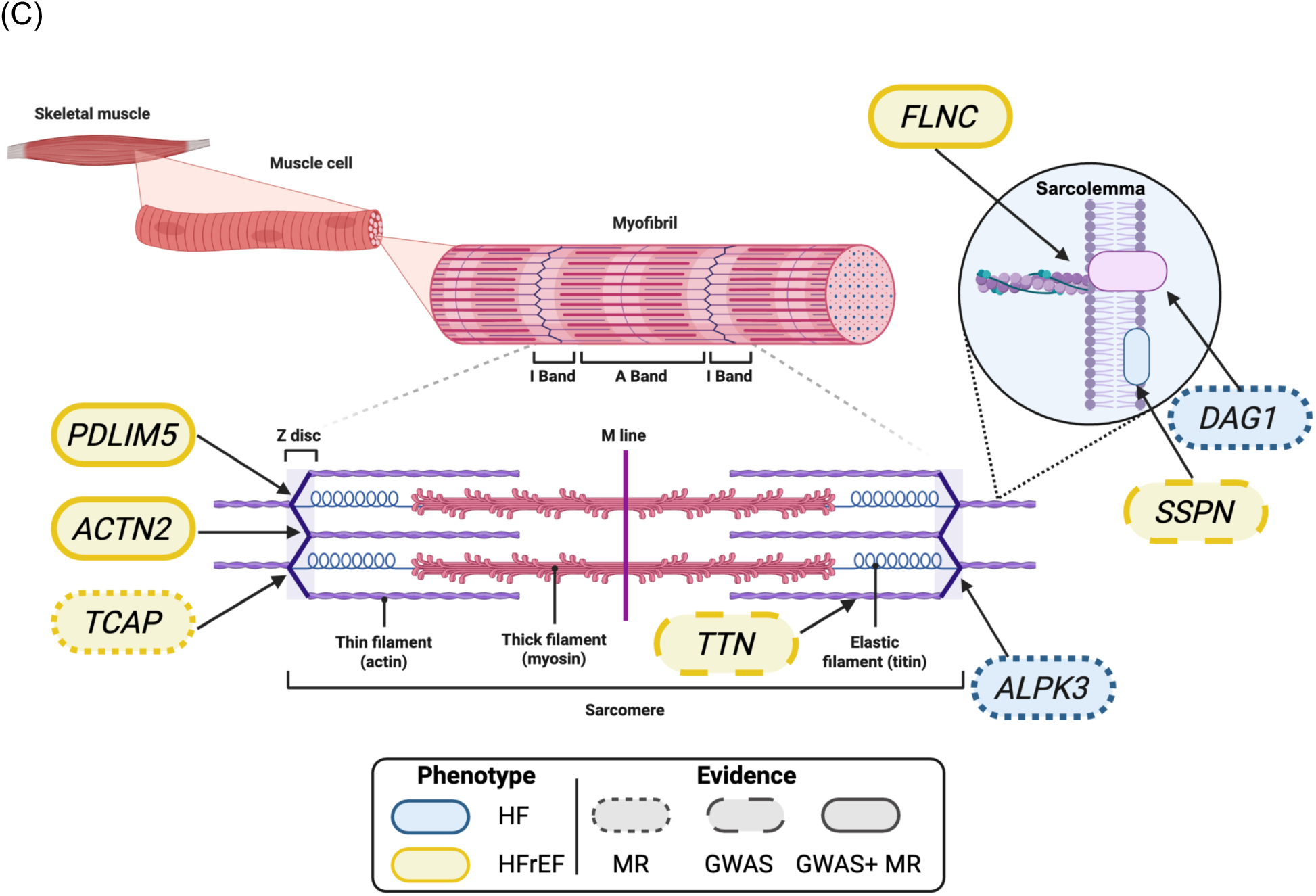
Cardiomyopathy annotations, and sarcomeric and sarcolemmal genes among HF-associated genes. (A) Venn diagrams showing the overlap of cardiomyopathy (CM) genes across HF, HFpEF, and HFrEF. The top panel displays all CM genes, the bottom left panel HCM genes, and the bottom right panel DCM genes. Numbers indicate genes shared across HF phenotypes. Only novel genes are annotated: blue indicates GWAS evidence, red indicates MR evidence, and green indicates support from both GWAS and MR analyses of HF, HFrEF, and/or HFpEF. Genes outlined in red are included in an Invitae DCM or HCM panel; all other genes were identified through DCM or HCM GWAS. (B) Dot plot of gene–trait associations for novel HF, HFrEF, and HFpEF findings across HF phenotypes, cardiac MRI traits, and orthogonal sources for HF/cardiomyopathy or cardiovascular evidence (OMIM/ClinGen, mouse knockout [KO] models, and secondary genes). HF and cardiac MRI associations are shown for GWAS (P < 5 × 10⁻⁸) in blue and MR (Bonferroni-significance) in red, where available, whereas orthogonal evidence is shown in green. For GWAS and MR results, point size reflects statistical significance (−log10(P)). For orthogonal evidence (green points), each point represents an evidence category (e.g., OMIM/ClinGen), and point size reflects the number of HF, cardiomyopathy, or cardiovascular evidence supporting the gene within that category. (C) Schematic of the muscle structure to the sarcomere and sarcolemma, annotated with genes for which GWAS and MR evidence support an association with HF and HF subtypes. Genes are positioned adjacent to their corresponding structural component (e.g. Z-disc, thin/thick filament, M-line), and color-coded by HF phenotype: blue indicates evidence for HF, and yellow indicates evidence for HFrEF. Border style denotes the type of supporting evidence: dotted outline indicates MR evidence only, dashed outline indicates GWAS evidence only, and solid outline indicates both GWAS and MR evidence. Created in BioRender. Rasooly, D. (2026) https://BioRender.com/2bzb36s *\*Cardiac MRI metrics with no significant associations are not displayed in the plot. These traits include left ventricular stroke volume (LVSV), right ventricular global longitudinal strain (RV-GLS), right ventricular global radial strain (RV-GRS), right ventricular end-diastolic volume (RVEDV), and mean global myocardial T1 (MGM T1)*. *\*\*Abbreviations. LVEDV, left ventricular end-diastolic volume; LVESV, left ventricular end-systolic volume; LVM, left ventricular mass; LVEF, left ventricular ejection fraction; LVGCS, left ventricular global circumferential strain (short axis); LV-GLS, left ventricular global longitudinal strain (long axis); LV-GRS, left ventricular global radial strain (short axis); RVESV, right ventricular end-systolic volume; and RV-GCS, right ventricular global circumferential strain (short axis). Traits indexed to body surface area are denoted by the suffix “i”*.

### Mechanistic pathways underlying genetic associations

#### Cardiovascular/Neurohormonal pathways

Five novel genes (*GUCY1A3*, *CYP11B2*, *ENPEP, PDE3A*, and *PTPRG*), and 3 known genes (*NOS3*, PCSK1, and *CASZ1*) are associated with effects on the vasculature as the main mechanism of action. *GUCY1A3*, *PDE3A*, and *NOS3* are all related to the nitric oxide (NO)-soluble guanylyl cyclase-cyclic GMP signaling pathway. *NOS3* encodes NO, which then binds a receptor encoded by *GUCY1A3*, and the *PDE3A* gene encodes an enzyme acting as an intracellular modulator of the response. *CYP11B2* (encodes aldosterone synthase) and *PCSK1* (an enzyme that uses renin as substrate) are novel genes involved in the renin-angiotensinogen aldosterone (RAAS) pathway. Additional genes in the RAAS pathway are *ENPEP*, which cleaves angiotensin II, and *CASZ1*, a mineralocorticoid regulator that regulates aldosterone production.

#### Hypothalamic leptin-melanocortin pathway

We identified 17 genes (7 novel, 2 reassigned and 8 known), which are known to be associated with monogenic disorders of obesity or are identified as strong genes for obesity using a rare-variant approach^14^. Three novel genes, *LEPR*, *SH2B1*, and *GNAS*, and four known genes, *MC4R*, *SIM1*, *PCSK1*, and *BDNF*, were mapped to the hypothalamic leptin-melanocortin pathway involved in food intake regulation (**Figure 3A**). To assess whether associations with HF phenotypes were driven by obesity, we compared the GWAS and MR beta coefficients for HF, HFrEF, and HFpEF against those for BMI, finding a strong correlation (r=0.84-0.85; **Figure 3B-C**).

**Figure 3.**
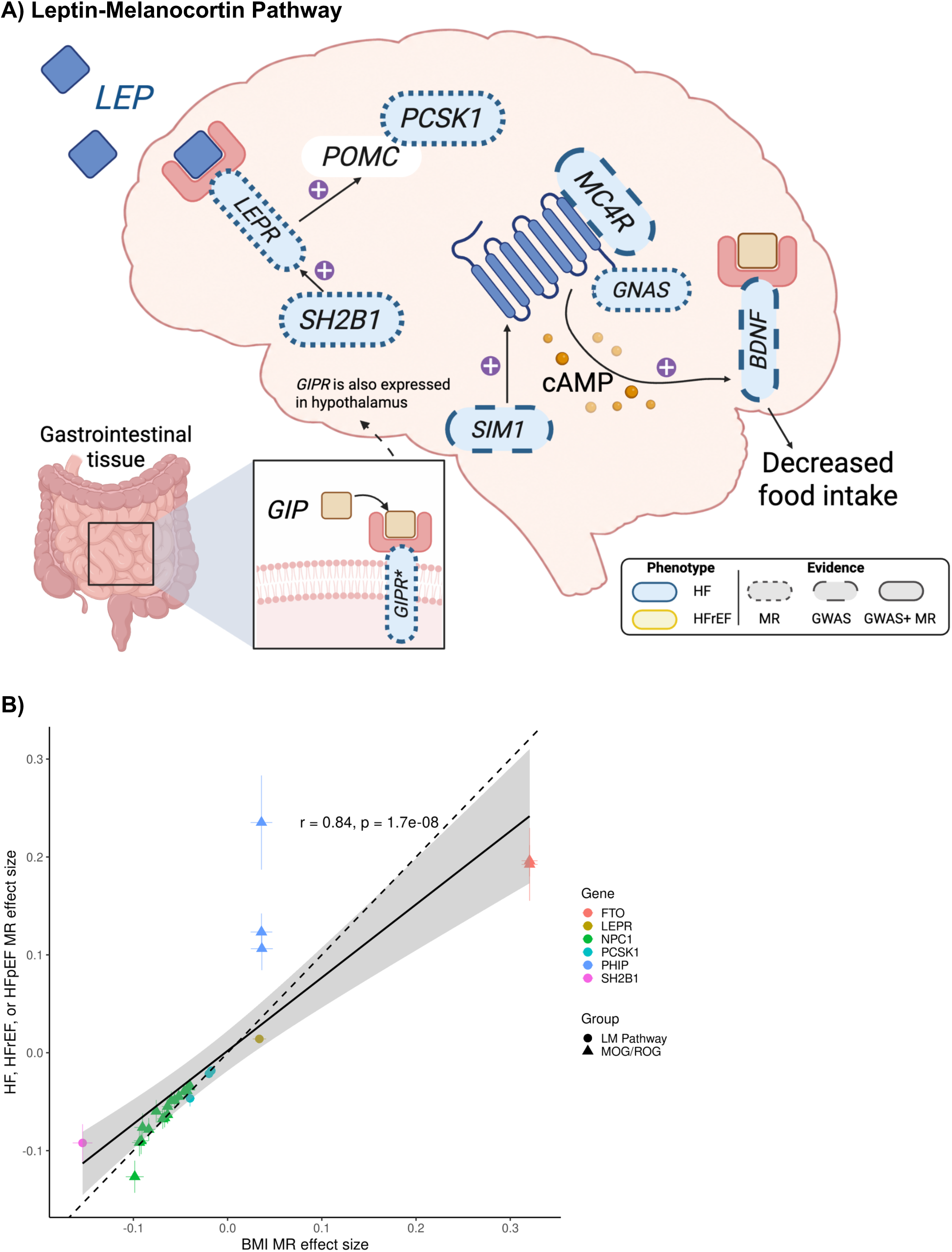
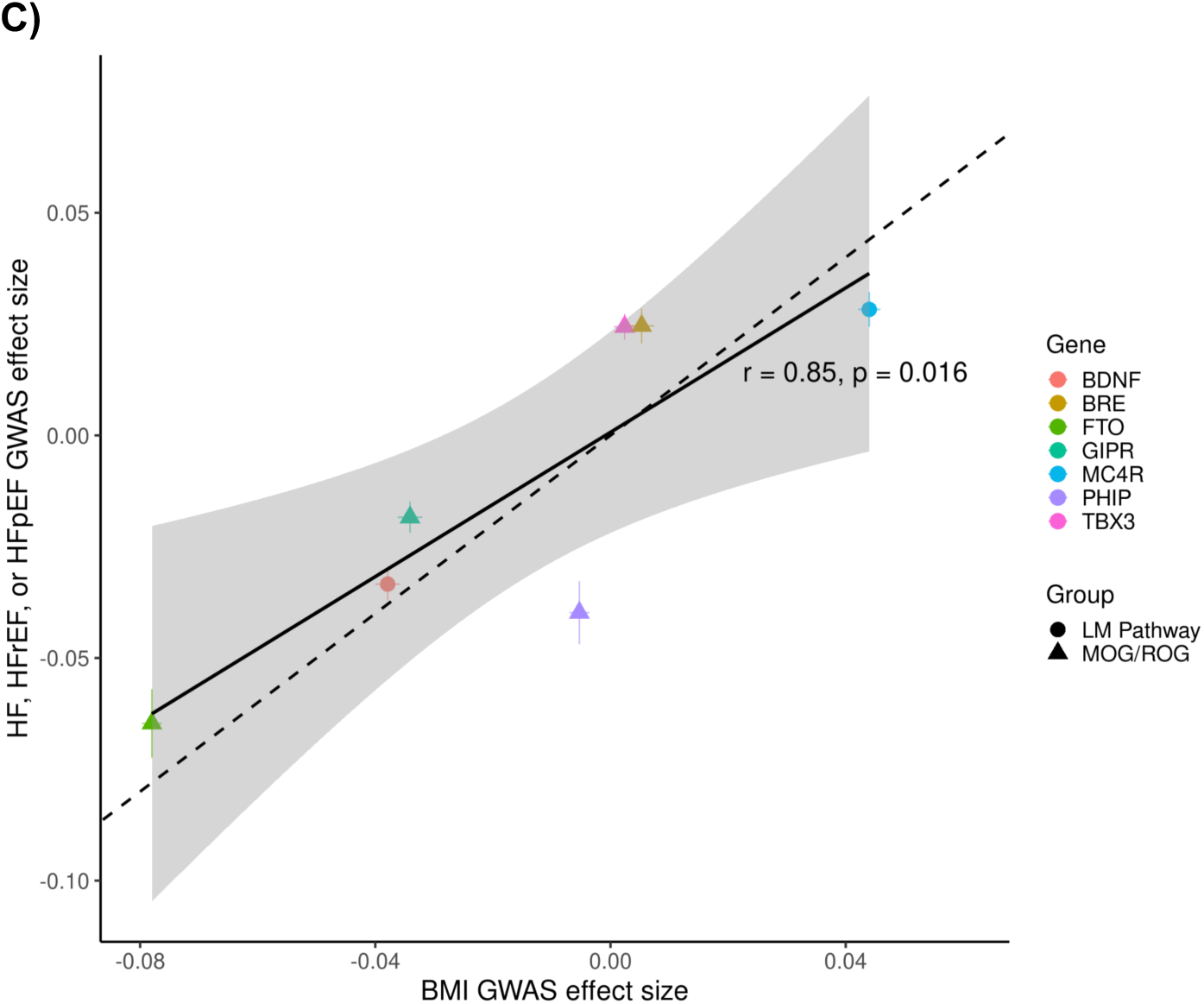
Leptin-melanocortin pathway and GIPR signaling implicated in obesity and heart failure. **(A)** Schematic of leptin-melanocortin signaling in the hypothalamus and its role in appetite regulation. Leptin (*LEP*) binds its receptor (LEPR) to stimulate *POMC* expression, which is processed by *PCSK1;* downstream melanocortin signaling through *MC4R* activates cAMP/*GNAS* signaling, leading to *BDNF* induction and decreased food intake. SH2B1 acts as a positive regulator of LEPR signaling. The lower inset represents GIP acting on GIPR in gastrointestinal tissue; GIPR is additionally expressed in the hypothalamus. Genes are colored blue to denote evidence for HF. Border style denotes evidence type: dotted outline indicates MR evidence only, dashed outline indicates GWAS evidence only, and solid outline indicates both GWAS and MR evidence. Asterisk denotes MR evidence significant at FDR < 5%. A plus sign with a purple background indicates a positive effect. Created in BioRender. Rasooly, D. (2026) https://BioRender.com/btsqm4b **(B)** Scatterplot comparing Mendelian randomization (MR) effect sizes for BMI (FDR < 5%) against HF and HF subtypes among gene-tissue pairs for monogenic and rare obesity genes (MOG/ROG) and leptin-melanocortin (LM) pathway genes. Each point represents a gene-tissue pair, colored by gene and shaped according to whether the gene belongs to the MOG/ROG or LM gene set. Horizontal and vertical error bars denote the standard error of the MR estimates. The solid black line indicates the fitted ordinary least squares regression across all points; the dashed line represents the identity line (y = x), corresponding to perfect concordance between BMI and HF-related effect sizes. The shaded band indicates the 95% confidence interval of the regression line. **(C)** As in (B), but comparing GWAS effect sizes for BMI against HF, HFrEF, and HFpEF.

#### Myocardial metabolism

We identified 24 genes associated with the metabolism of FAs (n=15), glucose (n=6), and branched-chain amino acids (BCAA, n=3), all sources of energy to the myocardium, while 56 genes were mapped to the mitochondria, a key organelle in energy production from FAs, glucose and BCAAs.

Genes related to triglyceride-rich lipoprotein-mediated lipolysis include *LPL*, *LIPC*, and *LMF1*. *LMF1* is required for the activation of *LPL*, and *LIPC* enzymes are involved in the lipolysis of circulating lipoproteins (**Figure 4A**). *LMF1* showed associations with measures of LV contractility (LV-GCS, and LV-GRS) and LV remodeling (LVEDV and LVESV).

**Figure 4.**
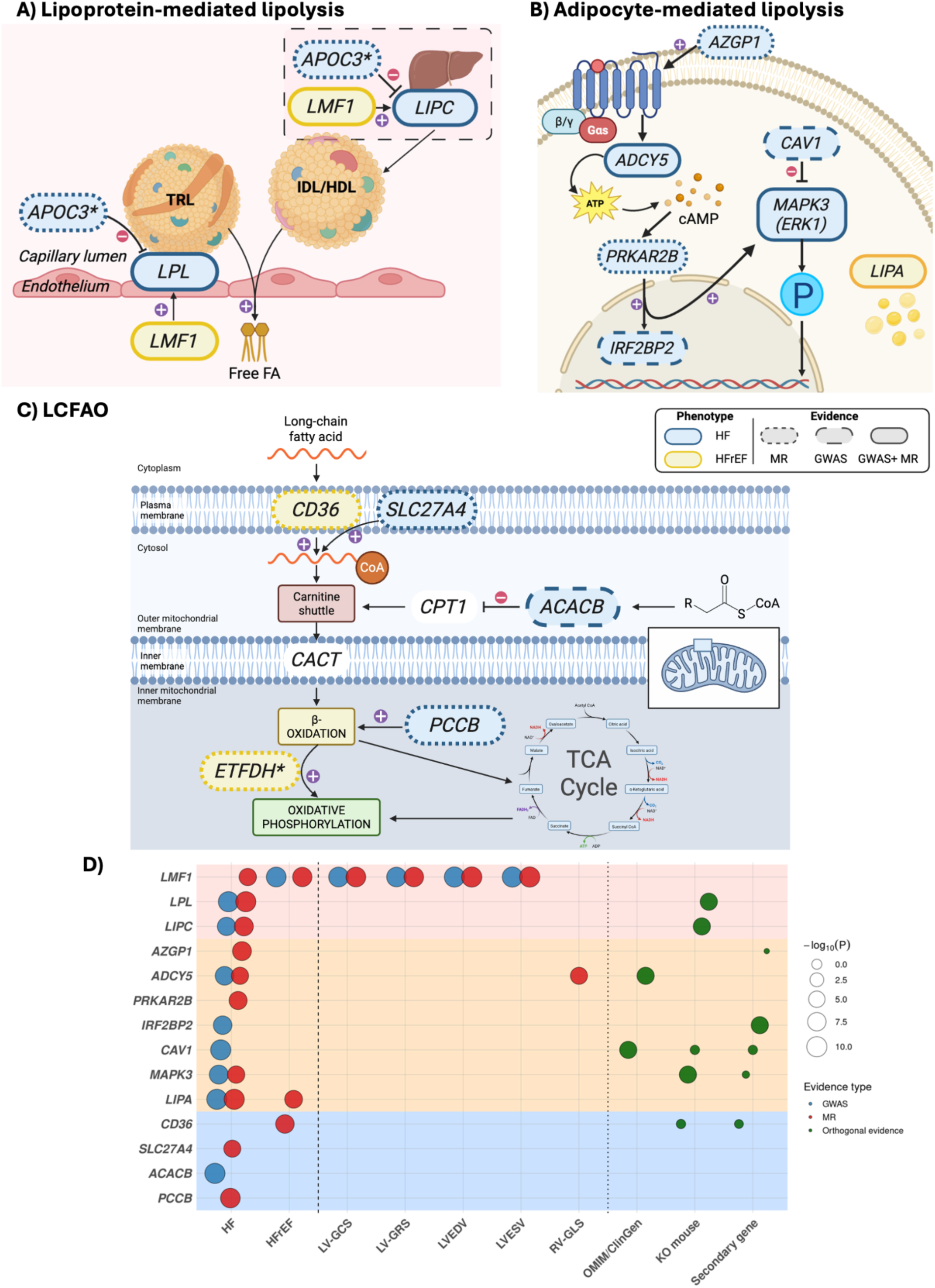
Lipoprotein-mediated and adipocyte-mediated lipolysis and long-chain fatty acid oxidation roles in HF. **(A)** Lipoprotein-mediated lipolysis and key HF-associated genes involved in this pathway are shown. LPL hydrolyzes triglycerides in very-low-density lipoproteins (VLDL) and chylomicrons, generating free fatty acids and monoacylglycerols. LMF1 is required for LPL maturation and activation, and for activation of LPL and LIPC. APOC3 inhibits LPL-mediated lipolysis. LIPC contributes to the release of free fatty acids from circulating lipoproteins, hydrolyzing triglycerides and phospholipids from LDLs and HDLs. Created in BioRender. Rasooly, D. (2026) https://BioRender.com/v473xxh **(B)** Adipocyte-mediated lipolysis highlighting key HF-associated genes is shown. AZGP1 modulates intracellular signaling through ADCY4 and cAMP, leading to activation of PRKAR2B and downstream phosphorylation events that regulate transcription, including IRF2BP2. PKA signaling cross-talks with the MAPK pathway, resulting in activation of MAPK3, which translocates to the nucleus to regulate gene expression. LIPA mediates lysosomal lipid degradation. CAV1 inhibits MAPK3 signaling. Created in BioRender. Rasooly, D. (2026) https://BioRender.com/5l5c7u6 **(C)** Long-chain fatty acid oxidation and key HF-associated genes involved in this pathway are shown. Long-chain fatty acids are imported through CD36 and converted to fatty acyl-CoA by SLC27A4. ACACB regulates mitochondrial fatty acid uptake by producing malonyl-CoA, an inhibitor of the carnitine shuttle. Fatty acyl-CoA is subsequently transported into mitochondria via the carnitine shuttle through CACT and undergoes β-oxidation, with electrons transferred to the respiratory chain through ETFDH. PCCB converts propionyl-CoA to succinyl-CoA, linking fatty acid metabolism to the tricarboxylic acid (TCA) cycle and oxidative phosphorylation. Across panels A-C, genes are colored blue to indicate evidence for HF and yellow to indicate evidence for HFrEF. Border style denotes the source of genetic evidence: dotted outlines indicate MR evidence only, dashed outlines indicate GWAS evidence only, and solid outlines indicate support from both GWAS and MR analyses. An asterisk denotes MR associations significant at FDR < 5%. Created in BioRender. Rasooly, D. (2026) https://BioRender.com/m7d2jy0 **(D)** Dot plot showing gene–trait associations across HF phenotypes, cardiac MRI traits, and orthogonal evidence for HF, cardiomyopathy, and cardiovascular disease (OMIM/ClinGen, mouse knockout [KO], and secondary gene evidence) for genes shown in panels A–C. Panels are color-coded to match the corresponding schematics (pink, lipoprotein-mediated lipolysis; cream, adipocyte-mediated lipolysis; blue, long-chain fatty acid metabolism). HF and cardiac MRI associations are shown for GWAS (blue; *P* < 5 × 10⁻⁸) and MR (red; Bonferroni-significant), where available, and orthogonal evidence is shown in green. For GWAS and MR results, point size reflects statistical significance (−log10[*P*]). For orthogonal evidence (green points), each point represents an evidence category (e.g., OMIM/ClinGen), and point size reflects the number of HF, cardiomyopathy, or cardiovascular evidence supporting the gene within that category. For genes with multiple MR associations, only the result with the smallest *P* value is displayed.

Genes related to adipocyte-mediated lipolysis include *AZGP1*, *ADCY5*, *PRKAR2B*, *CAV1*, *MAPK3*, *IRF2BP2*, and *LIPA*. *AZGP1* is known to stimulate lipolysis through the cAMP pathway, wherein *ADCY5* catalyzes cAMP production from ATP to activate protein kinase (PKA). *PRKAR2B* encodes the regulatory subunit beta of PKA, a primary intracellular mediator of hormonally stimulated lipolysis. *MAPK3*, also known as ERK1, is also involved in hormonally induced lipolysis downstream of PKA. *CAV1* facilitates PKA-mediated phosphorylation of perilipin. *IRF2BP2* suppresses lipolysis in adipocytes by inhibiting hormone-sensitive lipase, a key enzyme in adipocyte lipolysis, while *LIPA* is involved in non-canonical adipocyte lipolysis (**Figure 4B**).

Genes involved in long-chain FA (LCFA) oxidation were *CD36*, *SLC27A4*, *ACACB*, *ETFDH*, and *PCCB* (**Figure 4C**). *CD36* and *SLC27A4* are transporters of LCFA channeling FAs into intracellular metabolism. *ACACB* acts as a potent inhibitor of carnitine palmitoyltransferase 1, the rate-limiting enzyme responsible for the transport of LCFA into the mitochondria. *PCCB* encodes the beta subunit of the mitochondrial enzyme propionyl-CoA carboxylase, which catalyzes the final step of odd-chain FA oxidation. *ETFDH* couples LCFA oxidation to oxidative phosphorylation and ATP production.

Genes involved in BCAA metabolism were *BCKDK* (novel) and *BCKDHA* (known), both identified as HF genes, and *YBX3* (novel), identified as an MR HFrEF gene. *BCKDHA* is the active core subunit of the BCKDH complex, which drives the rate-limiting step of BCAA oxidation, while *BCKDK* inhibits this complex by phosphorylating BCKDHA and thereby controlling the breakdown of BCAAs. *YBX3* stabilizes *SLC3A2* mRNA (BCAA transporter) and hence promotes the uptake of BCAA into adipocytes, while also facilitating brown adipogenesis and thermogenesis.

Genes involved in glucose metabolism were *SLC2A4RG*, *SLC37A4*, *ENO3*, *VTI1A*, *BACH1*, and *PACSIN3*. *SLC2A4RG* is a transcription factor that activates the expression of GLUT4 in adipose and muscle tissues. VTI1A allows GLUT4 to move from intracellular storage sites to the plasma membrane after insulin stimulation, facilitating glucose uptake in adipose and muscle tissues. *SLC37A4* enables the hydrolysis of glucose-6-phosphate, with subsequent release of glucose into the bloodstream. *PACSIN3* regulates glucose metabolism in adipocytes by increasing trafficking and localization of GLUT1 to the plasma membrane, and ENO3 encodes beta-enolase, a key glycolytic enzyme primarily expressed in skeletal muscle. *BACH1* shifts metabolism from oxidative phosphorylation towards glycolysis. *BACH1* has 24 genes directly linked to it that were either reported as HF or CM GWAS genes or were supported by animal genetic evidence for HF.

#### Mitochondrial proteome

Overall, 42 novel, 4 reassigned and 10 known HF, HFrEF, and HFpEF genes were mapped to the mitochondrial proteome based on human mitochondrial proteome annotations integrating experimentally supported and computationally predicted mitochondrial localization (**Table S20**). Of the 56 genes, 13 (23%) had support from at least one orthogonal source. Most genes map to the metabolism of lipids, including LCFA (n=9), amino acids (n=5), oxidative phosphorylation (n=9), and mitochondrial central dogma (n=10).

#### Adipokines

We identified 22 genes (14 novel, 7 known, and 1 reassignment) that encode proteins considered as adipokines (**Table S21**). The proposed mechanisms for the 10 novel genes included inflammation (*CD14*, *IL6*, *TNFS13*, *ERAP1*), lipolysis (*AZGP1*), thermogenesis (*SLIT2* and *CRLF1*), fibrosis (*AZGP1*) and angiogenesis (*VEGFB*). The biological roles of the remaining adipokine genes are described in **Table S21**, including additional genes classified as adipokines that passed FDR 5% in our MR analysis.

#### Neuregulin-ERBB and activin pathways

We found multiple genes associated with the neuregulin-ERBB and activin signaling pathways. Within the neuregulin-ERBB pathway, a recognized protective system in HF, we identified the ligands *NRG1* and *HBEGF*, receptor *ERRB2*, and the intracellular signaling component *MAPK3*. At FDR 5%, we identified the receptors *ERBB4* and *ERBB3* (**Figure 5A**). Within the activin pathway, which regulates muscle growth and cardiac remodeling, we identified the receptors *ACVR2A* and *TGFBR1*, the intracellular signaling mediator *SMAD3*, and the regulators *FSTL3* and *SMAD7*. *FSTL3* inhibits the ligands activin A (*INHBA*), a target of sotatercept (approved for pulmonary hypertension), and *GDF11*; both genes were suggestive findings for HF at FDR 5% (**Figure 5B,C**).

**Figure 5.**
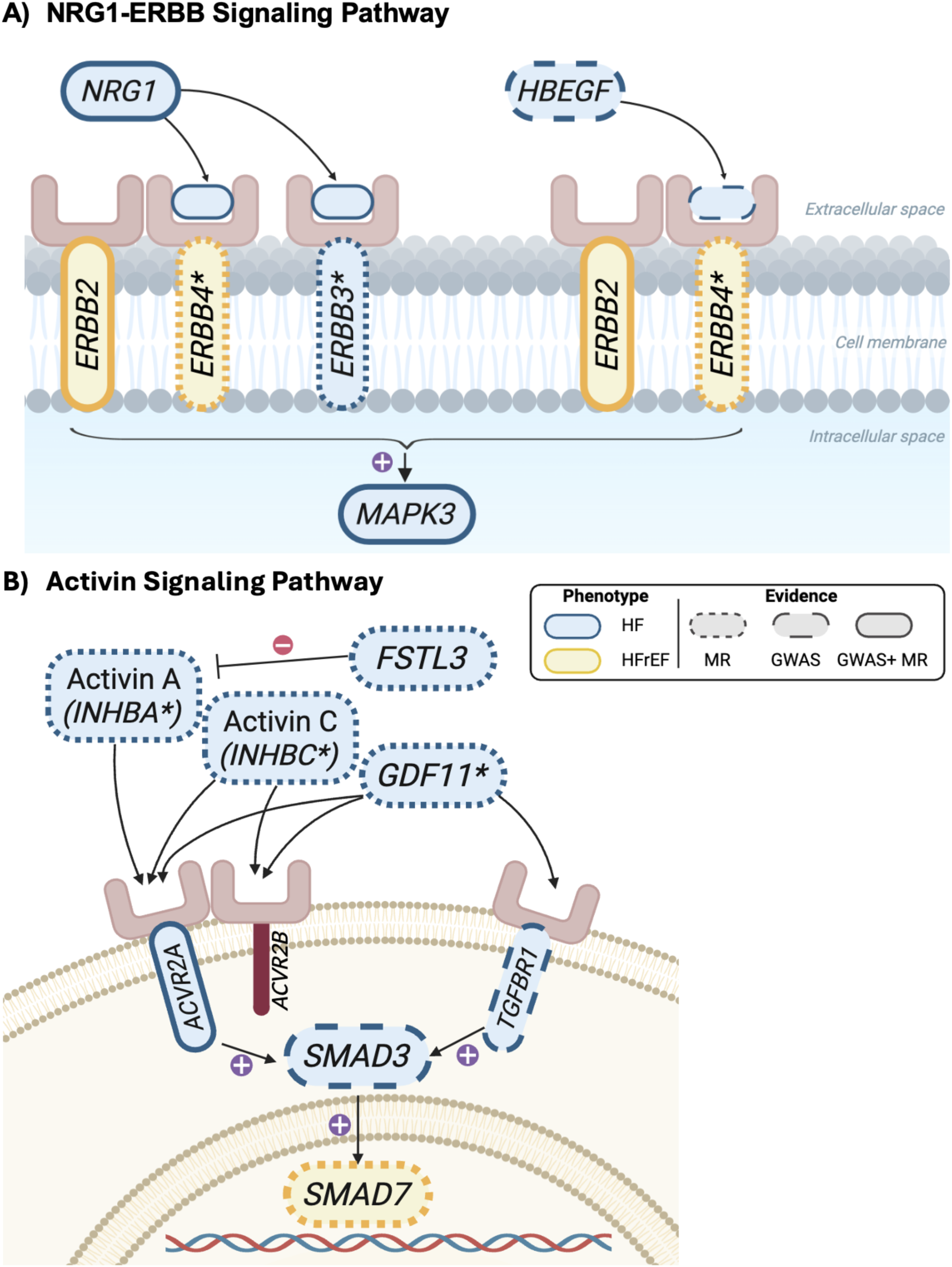
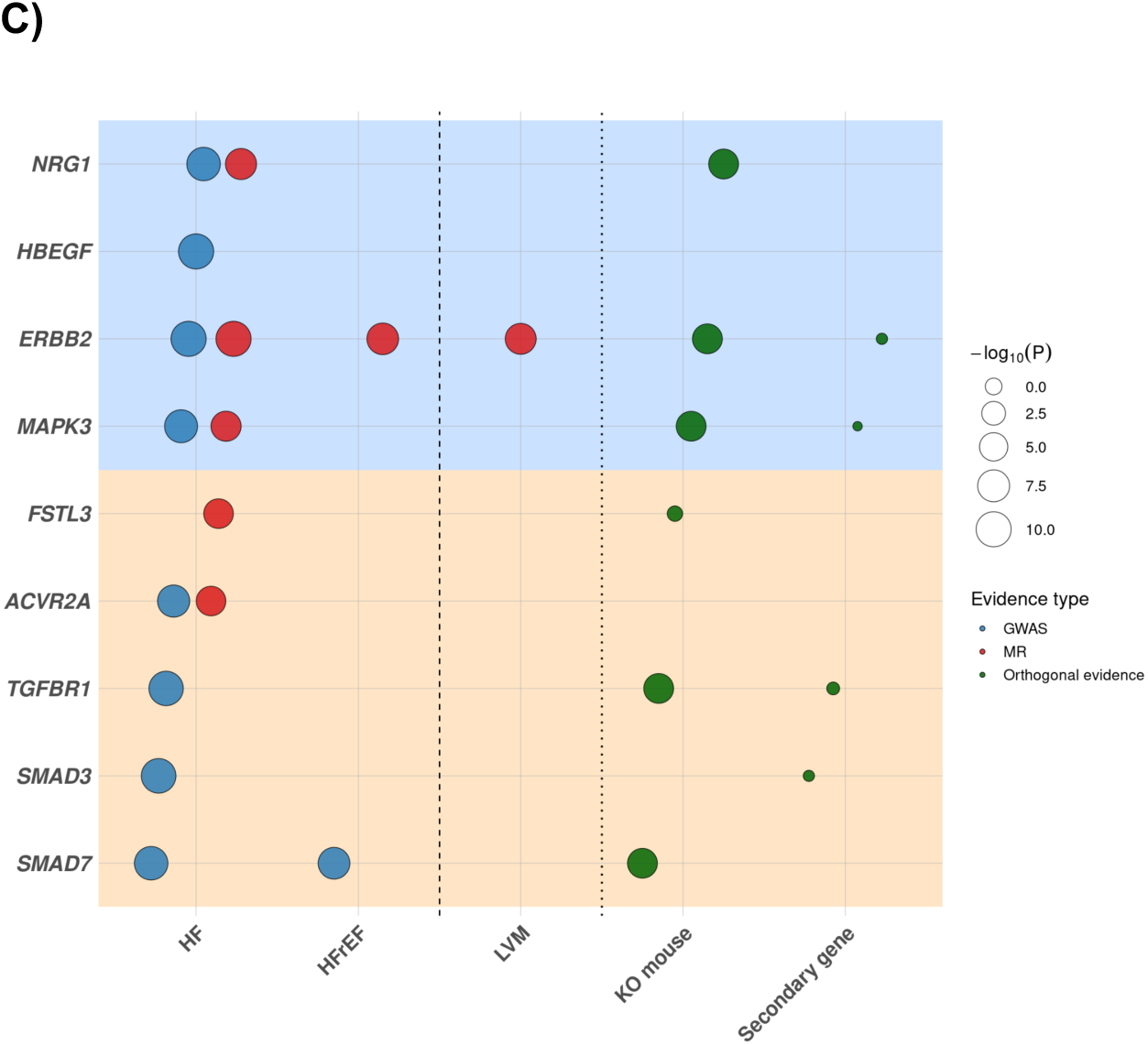
NRG1-ERBB and Activin Signaling Pathways Implicated in HF. (A) Schematic of NRG1 and HBEGF-ERBB signaling at the cell membrane. NRG1 is a novel HF-associated gene encoding a ligand that signals through its co-receptors ERBB4 and ERBB3, typically via heterodimerization with ERBB2. ERBB4 represents a novel association with HF and HFrEF, while ERBB3 is a previously reported HF association. ERBB2 functions as a co-receptor to ERBB4 and is supported by multiple lines of genetic evidence. HBEGF is a known HF association encoding a ligand for EGFR and ERBB4, with downstream signaling often involving ERBB2-containing receptor dimers. NRG1 and HBEGF signaling converge on activation of the mitogen-activated protein kinase (MAPK)/extracellular signal-regulated kinase (ERK1/2) cascade, including MAPK3, which regulates cellular proliferation, survival, and stress. In Panels A and B, Genes are colored to denote HF phenotype evidence — yellow for HFrEF, blue for HF — and bordered to denote evidence type: solid outline indicates GWAS and MR evidence, dotted outline indicates MR evidence only, and dashed outline indicates GWAS evidence only. Asterisk denotes MR evidence significant at FDR < 5%. Created in BioRender. Rasooly, D. (2026) https://BioRender.com/boc7x42 **(B)** Schematic of activin type II receptor signaling. Activin A (encoded by *INHBA*, a novel HF gene) and activin C (encoded by *INHBC*, a novel HF gene), as well as *GDF11*, function as ligands for the activin type II co-receptors *ACVR2A* (a novel HF gene) and *ACVR2B*. Ligand binding to *ACVR2A/B* leads to phosphorylation of *SMAD3* (a novel HF gene), with downstream transcriptional responses in the nucleus involving *SMAD7* (a novel HF gene). Created in BioRender. Rasooly, D. (2026) https://BioRender.com/gdswcdv **(C)** Dot plot showing gene–trait associations across HF phenotypes, cardiac MRI traits, and orthogonal evidence for HF, cardiomyopathy, and cardiovascular disease (OMIM/ClinGen, mouse knockout [KO], and secondary gene evidence) for genes shown in panels A and B. Panels are color-coded to match the corresponding schematics (blue, NRG1-ERBB pathway; cream, activin signaling pathway). HF and cardiac MRI associations are shown for GWAS (blue; *P* < 5 × 10⁻⁸) and MR (red; Bonferroni-significant), where available, and orthogonal evidence is shown in green. For GWAS and MR results, point size reflects statistical significance (−log10[*P*]). For orthogonal evidence (green points), each point represents an evidence category (e.g., OMIM/ClinGen), and point size reflects the number of HF, cardiomyopathy, or cardiovascular evidence supporting the gene within that category. For genes with multiple MR associations, only the result with the smallest *P* value is displayed.

## DISCUSSION

In the largest multi-ancestry genetic study to date of HF and its major clinical subtypes, we identified 166 novel loci and 375 novel putatively causal genes associated with HF, HFrEF, and HFpEF. The large catalog of novel and known genes allowed us to resolve, at high resolution, a landscape of HF-relevant pathways with therapeutic potential. Our findings on various interconnected metabolic pathways and adipokines suggest that adipose tissue should be considered as a key target of future interventions for HF, and in particular HFpEF^15^, prevention and management.

We identified multiple genes associated with cardiomyocyte contraction such as components of the sarcomere Z-disc, M-band, and dystrophin-glycoprotein complex. In addition, we validated targets of therapies currently approved for the management of different stages of HF (*GUCY1A1*/Vericiguat, *ATP1B2*/Digoxin, *PDE3A*/Milrinone), retroactively confirming the value of human genetics for drug target discovery. Furthermore, we validated targets of JK07 (*NRG1*) and Baxdrostat (*CYP11B2*), which are currently under evaluation for HFrEF and HFpEF^16^ and for the primary prevention of HF^17^, respectively. Lastly, we identified potential repurposing opportunities for HF involving type-II activin receptor antagonist (*ACVR2A*)^18,19^ and analogues of human leptin (*LEPR*)^20^. This adds to the growing list of genetically-supported cardiometabolic drug targets (*IL6*, *LPA*, *ADM*, *PCSK9*, *ENPEP*, *CACNB2*, *EDNRA*, *MC4R*, and *RXFP2*). Therefore, our results that span the spectrum from known and investigational therapies to repurposing opportunities demonstrate the power of genetically-informed approaches to drug development in HF.

Interestingly, we mapped nearly 100 genes for HF, HFrEF, and HFpEF to interconnected metabolic pathways relevant to HF, such as food intake, lipolysis, metabolism of FAs, BCAA, glucose, and the mitochondrial proteome which is involved in energy production. Our genetic findings on the leptin-melanocortin pathway further corroborates a pathway that is supported by various randomized trials of incretin-based therapies, which have demonstrated benefits in HFpEF patients, while results in HFrEF remain inconclusive^21^. However, our findings of six genes associated with adipocyte-driven lipolysis raise the profile of this mechanism as a therapeutic pathway for HF in addition to weight loss. For example, *PRKAR2B* is highly expressed in adipose tissue and its disruption in rodent models is associated with a lean phenotype, increased lipolysis, and elevated metabolic rate; our findings on HF suggest that this could be a dual target for weight loss and HF management.

Lipolysis, either from the adipocyte or lipoprotein-mediated, is the main mechanism for the delivery of free LCFAs to tissues with high energetic demand, such as the myocardium, especially in HF patients. *LMF1*, *LPL*, *MAPK3*, and *AZGP1* (involved in lipolysis) and *CD36* and *SLC27A4* (involved in the internalization of LCFA for further beta oxidation), all supported by MR, showed that an increase in gene expression or protein levels is associated with HF risk reduction. This is consistent with animal genetic studies showing that cardiac-specific overexpression of *CD36*, *CPT1*, *ACSL1*, and *ACADL*, which increase LCFAO, and cardiac-specific inhibition of *ACC2,* which increases LCFAO, are protective against HF^22–26^.

Further, human genetic evidence supporting the relevance of cardiac energetics to HF comes from genes associated with BCAA and glucose metabolism. We identified *BCKDK*, a novel HF gene that is highly expressed in cardiac tissue that inhibits the BCKDH enzyme complex that catalyzes the irreversible step in BCAA catabolism. Interestingly, the alpha subunit of the BCKDH complex is encoded by the *BCKDHA* gene, which is also highly expressed in cardiac tissue, and is previously reported and replicated by us as a GWAS HF gene. In agreement with our genetic findings, multiple studies have shown that pharmacological inhibition of *BCKDK*, which increases BCAA catabolism, leads to improvement in cardiac function in various rodent HF models^27,28^. Regarding glucose metabolism, we identified *ENO3* and *SCL37A4*, which are directly involved in glucose processing; *SLC2A4RG* and *BACH1*, which are involved as regulators of glucose metabolism; and *VTI1A* and *PACSIN3*, which are involved in the trafficking of *GLUT1* and *GLUT4* receptors to the plasma membrane. All these genes were highly expressed in skeletal muscle, liver, and adipose tissues, further highlighting the relevance of key metabolic tissues as therapeutic targets for HF management.

The neuregulin-ERBB and the activin signaling pathways were two mechanisms in which we identified multiple genes encoding ligands, receptors, and intracellular mediators of these pathways. Our main finding on the role of neuregulin-ERRB pathway in HF was the association of anti-ErbB2 therapy (used in breast cancer) with cardiac dysfunction as an adverse event, suggesting that agonism of this pathway may be beneficial in HF. Recently, JK07, an agonist of *ERBB4* currently in Ph2a study in HFrEF and HFpEF, was tested in a first-in-human study in HFrEF patients with encouraging results^29^. Similarly, the activin pathway has multiple investigational drugs, such as Bimagrumab and Sotatercept^30^, that could be repurposed for HF management based on our genetic findings.

We noticed that activin A and NRG4, ligands of the above pathways, have been described as key adipokines with a role in HFpEF^15^. This prompted us to revisit our findings with subsequent identification of more than 20 genes, mostly HF-associated, whose soluble proteins can be considered as adipokines, with paracrine or endocrine actions relevant to HFpEF, including inflammation, fibrosis, and angiogenesis, in addition to the lipolysis and thermogenesis mechanisms already described.

From our novel HFpEF genes, we identified through GWAS, the rs533557749/*RUNX1* locus that has various lines of evidence supporting our HFpEF findings. First, *RUNX1* is upregulated in cardiac tissue of patients with HCM compared with controls^31^. Second, inhibition of *RUNX1* either by genetic deletion in cardiomyocytes, RNA interference using adeno-associated virus serotype 9, or through pharmacological inhibition, showed benefit in a mouse HFpEF model^32,33^. Third, fibroblast-specific– but not cardiomyocyte-specific– deletion of *RUNX1* was found to be protective against adverse cardiac remodeling following myocardial infarction^34,35^. Interestingly, we noticed that *CDK6*, a MR hit for HF that we replicated, is a known regulator of *RUNX1* expression. *CDK6* genetic deletion in mice has shown an increase in beige adipose tissue formation and enhanced energy expenditure, findings that appeared mediated by *RUNX1*^36^. However, we note that this finding was driven by the MVP Asian American cohort and warrants replication.

Our study has important limitations that highlight directions for future research. First, our novel findings for HFrEF and HFpEF require replication in additional, adequately powered studies with access to these phenotypes, which are currently limited. Second, despite substantial efforts to increase genetic diversity, European ancestry remains the predominant source of evidence, accounting for 67% to 85% of participants. Third, locus independence was defined using ±250kb genomic distance, a common GWAS convention that does not account for long-range LD. Fourth, as is typical in GWAS, assigning genetic loci to their causal genes remains a substantial challenge. To address this, we employed four complementary gene-mapping strategies and incorporated MR-QTL analysis as an orthogonal strategy for discovery. In addition, we emphasized the interpretation of our findings on mechanisms, rather than single genes. Given that multiple genes within a single mechanism are located on different chromosomes, we minimize the risk of spurious findings due to LD. Finally, many identified mechanisms were interconnected with support beyond human genetics.

In conclusion, we have substantially increased the number of loci and genes associated with HF, HFrEF, and HFpEF. We identified repurposing opportunities for existing investigational CV drugs and multiple novel genes within interconnected metabolic pathways relevant to HF, highlighting potential therapeutic targets for HF prevention and management.

## METHODS

The Million Veteran Program (MVP) is approved by the Department of Veterans Affairs Central Institutional Review Board (IRB). All participants provided written informed consent. This study was conducted in accordance with the principles outlined in the Declaration of Helsinki and has received ethical and study protocol approval by the IRB.

### Study population

The HF meta-analysis included 4,468,166 individuals, of whom 345,687 were HF cases and 4,122,479 were control individuals. The HFpEF meta-analysis included 1,514,078 individuals, including 47,192 cases and 1,466,886 control individuals. The HFrEF meta-analysis included 1,695,762 individuals, including 46,934 cases and 1,648,828 control individuals. Individuals were drawn from the Million Veteran Program (MVP)^37^, HERMES Consortium^6^, Biobank Japan^38^, the Global Biobank Meta-analysis Initiative^39^, the All of Us Research Program^40^, the Mexico City Prospective Study^41^, and FinnGen Release 12^42^. The study population included individuals of European, African, Asian, and Admixed American ancestry. All contributing studies received IRB approval and obtained informed consent.

### Million Veteran Program

The Million Veteran Program (MVP) is a large, voluntary research program embedded within the U.S. Department of Veterans Affairs (VA) health care system, integrating genomic data with longitudinal electronic health record (EHR) data, including diagnostic codes, laboratory measurements, and imaging^37^.

HF cases within the MVP were identified using International Classification of Diseases (ICD)-9 code 428.x and ICD-10 code I50.x in conjunction with echocardiographic data obtained within six months of diagnosis. Accurate subclassification into HFpEF and HFrEF requires reliable capture of left ventricular ejection fraction (LVEF), which is frequently missing or fragmented across structured EHR fields. To overcome this, we applied a validated natural language processing (NLP) pipeline that extracted LVEF values from both structured and unstructured electronic health record (EHR) data, including echocardiogram reports, nuclear medicine reports, cardiac catheterization reports, history and physical examination notes, progress notes, discharge summary notes, and other clinical notes– capturing LVEF values measured outside the VA system and thereby reducing HF misclassification due to incomplete LVEF ascertainment^43^.This comprehensive LVEF capture ensured that we obtained the LVEF measured at the time of HF diagnosis, ensuring accurate HFpEF classification and excluding veterans with recovered LVEF from the HFpEF cohort. Individuals were classified as HFpEF if the first recorded LVEF was ≥50% and as HFrEF if LVEF ≤40%. The phenotyping algorithm, which also incorporated B-type natriuretic peptide (BNP) testing to strengthen HF case ascertainment, achieved a 96% positive predictive value against blinded physician review^44^. Cohort curation and validation have been described previously^44^ and this phenotyping approach has been validated and applied across multiple epidemiological and genomic studies^45–47^. Genetic ancestry (European, African American, Asian American, or Hispanic) was assigned using self-report and principal components of ancestry, as previously described^73^.

Genotyping was performed using the MVP 1.0 custom Axiom array, developed as a single assay to be used on the multi-ethnic MVP cohort^48^. Standard quality control excluded duplicate or related samples, call rate <98.5%, and sex discrepancies. Imputation was conducted with Minimac4 using the 1000 Genomes Project Phase 3 v5 reference panel. Variants with minor allele frequency >1% were tested for association using PLINK2, adjusting for age, sex, and the first ten genetic principal components.

### HERMES Consortium

We obtained GWAS summary statistics from the HERMES Consortium, comprising 1,946,349 individuals across 42 studies^6^. HF phenotypes were harmonized across studies using a standardized framework, including overall HF, non-ischemic HFrEF (LVEF <50%), and non-ischemic HFpEF (LVEF ≥50%), based on physician diagnosis, hospital records, and/or diagnostic codes, with LVEF derived from cardiac imaging where available. Study-level genome-wide association analyses were performed using logistic regression for prevalent outcomes or Cox proportional hazards models for incident outcomes under an additive genetic model, adjusting for age, sex, genetic principal components, and study-specific covariates. Summary statistics were subsequently combined using fixed-effect inverse-variance weighted meta-analysis implemented in METAL. Further details on phenotype harmonization, genotyping, and quality control procedures have been described previously^6^.

### Global Biobank Meta-analysis Initiative

The Global Biobank Meta-analysis Initiative (GBMI) conducted a large-scale, multi-ancestry HF GWAS leveraging genetic and electronic health record data from 21 biobanks across four continents^39^. The discovery analysis, excluding UK Biobank, included 60,605 HF cases and nearly 860,000 control individuals across six ancestral populations. HF was defined using ICD-based phecodes, without separation into clinical subtypes. We used GBMI summary statistics in our HF meta-analysis.

### FinnGen R12

We obtained GWAS summary statistics from FinnGen Release 12^42^, including data on 38,165 HF cases of European ancestry. HF was defined using harmonized national ICD-9 and ICD-10 codes derived from Finnish national health registries. Genotyping was performed using genome-wide arrays and imputed to the SISu v4.2 imputation reference panel. Association analyses were conducted using a logistic mixed-model framework under an additive genetic model in Regenie (version 2.2.4), adjusting for age, sex, batch effects, top 10 genetic principal components, Finngen chip version (1 or 2), and legacy genotyping batch. Further details on study design, phenotype definition, and analysis have been described previously^42^.

### Biobank Japan

For the HFrEF and HFpEF analyses only, we obtained GWAS summary statistics from Biobank Japan^38^. In Biobank Japan, HF subtypes were defined based on physician diagnosis and LVEF, with HFrEF defined as LVEF <40% and HFpEF as LVEF >50%. Genome-wide association analyses were performed using Regenie under an additive logistic regression model, adjusting for age, age squared, sex, and the top 10 genetic principal components. Genotypes were array-based and imputed to a population-specific Japanese reference panel, with variants filtered at minor allele frequency (MAF) ≥ 0.01 and imputation quality score (INFO) > 0.3 prior to analysis. Further details regarding study design, genotyping, imputation, and quality control procedures have been described previously^38^.

### All of Us Research Program

In the All of Us Research Program^40^, HF cases were identified using electronic health record data mapped to the OMOP common data model. Individuals were classified as HF cases if they had ≥2 occurrences of ICD-9-CM or ICD-10-CM diagnosis codes for HF recorded on distinct dates, including ICD-9-CM codes 398.91 and 428.x and ICD-10-CM codes included I09.81 and I50.x (including all subcategories). Individuals with only a single recorded HF diagnosis code were excluded to improve specificity, and individuals not meeting HF diagnosis criteria were classified as control individuals. Genome-wide association analyses were performed under an additive genetic model using PLINK v2.0, adjusting for age, sex, and the first 20 genetic principal components to account for population structure. Variants were filtered to include those with minor allele frequency (MAF) ≥ 0.01 and imputation quality score (INFO) ≥ 0.3. Standard quality control excluded variants with Hardy–Weinberg equilibrium *P* < 1×10⁻⁶ in controls and call rate <98.5%, and excluded samples with call rate <98.5%, sex discordance, or excess relatedness. Analyses were performed in a combined multi-ancestry sample, and association results were reported as odds ratios with corresponding standard errors and *P* values.

### Mexico City Prospective Study

In the Mexico City Prospective Study (MCPS), HF cases were defined using ICD-10 codes linked to mortality records adjudicated by MCPS investigators. Cases comprised 1,219 deceased individuals whose adjudicated primary cause of death was classified as chronic rheumatic heart disease (I05–I09), hypertensive heart disease (I11), chronic ischaemic heart disease (I25), non-rheumatic valvular disorders (I34–I36), cardiomyopathy (I42), or HF (I50). Controls included 136,285 individuals who were alive as of January 1, 2021 and had no record of these conditions, excluding individuals with uncertain cause of death or a self-reported diagnosis of coronary heart disease at recruitment. Genotyping was performed using the Illumina Global Screening Array with previously described methods for quality control and imputed to the TOPMed reference panel (version r2)^41^. Variants with imputation quality r² > 0.4 and effective sample size (N_eff) > 50 were retained^49^, yielding 7,375,232 variants for analysis. Genome-wide association analyses were conducted using Regenie v3.1.3 under a logistic regression framework, adjusting for age, age squared, sex, and the first seven genetic principal components. After excluding individuals with missing covariate data, 1,210 cases and 135,727 controls were included in the final analysis.

### Genome-wide association meta-analysis of HF, HFrEF, and HFpEF

We conducted inverse variance–weighted fixed-effects meta-analyses of genome-wide association summary statistics using METAL (v2020-05-05)^50^. Analyses included seven cohorts across four genetic ancestry groups (European, Asian, African and Admixed American). Variants were retained if minor allele frequency (MAF) was >1% and imputation quality (INFO) >0.3 in contributing studies. Variants in the meta-analysis were aligned to the same effect allele, and ambiguous or poorly imputed variants were excluded.

Between-study heterogeneity was assessed in METAL. A total of 13,843 FinnGen cases and 121,422 controls overlapped between HERMES (which includes FinnGen R3) and FinnGen R12. To account for this sample overlap, *P* values were calculated using a sample size–weighted approach incorporating study-specific sample size and effect direction, and effect estimates and standard errors were obtained using inverse variance–weighted meta-analysis. Inflation factors were calculated as the median of the observed test statistics divided by the expected median under the null hypothesis.

### Genomic risk loci definition and gene annotation

Genomic risk loci were defined using FUMA (v1.8.0) with the 1000 Genomes Project Phase 3 multi-ancestry reference panel^51^. Independent significant variants were identified among genome-wide significant SNPs (*P* ≤ 5 × 10⁻⁸) using linkage disequilibrium (LD)–based clumping (r² ≥ 0.6). Lead SNPs were defined as independent significant variants in low LD with one another (r² < 0.1). Genomic risk loci were constructed by merging LD blocks (r² ≥ 0.6) located within 250 kb of one another. For gene annotation, lead SNPs were mapped to the nearest gene based on physical distance to the transcription start site, using a 10 kb positional mapping window. Functional consequences of variants were annotated using ANNOVAR.

Gene-level association analyses were conducted using MAGMA (v1.10)^51^. SNPs were mapped to genes based on the meta-analysis summary statistics, using a window of 35 kb upstream and 10 kb downstream of each gene. MAGMA performs gene-based association tests using a multiple regression framework that models the joint effects of SNPs within a gene while accounting for LD.

### Functional annotation of variants

Genome-wide significant variants were functionally annotated using the Ensembl Variant Effect Predictor (VEP) to determine predicted molecular consequences and affected feature types^52^. Coding variants were further evaluated using in silico prediction tools. Deleteriousness of missense variants was assessed using SIFT (deleterious if <0.05) and PolyPhen-2 (“possibly damaging” or “probably damaging”). AlphaMissense scores were used to classify variants as likely pathogenic (>0.564), ambiguous (0.34–0.564), or likely benign (<0.34). REVEL scores (≥0.5) were used to prioritize missense variants with higher predicted pathogenicity. Combined Annotation Dependent Depletion (CADD) scores were used to quantify deleteriousness, with scores ≥20 considered indicative of high functional impact.

### Gene Mapping Methods

Candidate genes at GWAS loci were prioritized using four complementary approaches: (1) cS2G (combined SNP-to-gene), a heritability-informed framework that integrates multiple SNP-to-gene linking strategies^53^; (2) V2G (variant-to-gene), which integrates functional annotations including eQTLs, chromatin interactions and regulatory element data to link variants to target genes^54^; (3) nearest gene, which assigns each variant to the nearest protein-coding gene based on genomic distance^51^; and (4) MAGMA gene-based association analysis, which aggregates SNP-level associations within gene boundaries to generate gene-level *P* values^51^.

For each locus, genes implicated by all four approaches were considered. The primary assigned gene was defined as the gene most frequently prioritized across methods; when ties occurred, all tied genes were retained.

### Credible set analysis of causal variants

Each independently associated sentinel SNP was fine-mapped separately under a single-causal-variant Bayesian framework. For each sentinel SNP, candidate variants within a ±500 kb window were retrieved using LDlinkR, based on the pooled 1000 Genomes Project reference populations and GRCh37 coordinates^55^. Variants returned by LDproxy were matched to the full GWAS summary statistics by genomic position and alleles. Bayesian fine-mapping was performed using the corrcoverage R package to estimate posterior probabilities for candidate variants. Variants were ranked in descending order of posterior probability, and 90% and 95% Bayesian credible sets were constructed by sequentially including variants until their cumulative posterior probability reached or exceeded 0.90 and 0.95, respectively^56^.

### Cis-acting protein quantitative trait loci (pQTL)

Cis-acting protein quantitative trait loci (cis-pQTLs) were obtained from GWAS summary statistics from the Fenland^57^, deCODE^58^, ARIC^59^ and UK Biobank Olink studies^60^. Fenland, deCODE and ARIC quantified plasma proteins using the SOMAscan v4 platform in predominantly European-ancestry populations, whereas UK Biobank used the Olink platform. The deCODE study included genetic association data for 4,907 proteins measured in 35,559 individuals; Fenland for 3,892 proteins in 10,708 individuals; and ARIC for 2,004 proteins in 7,213 European Americans and 1,618 proteins in 1,871 African Americans. The UK Biobank Olink dataset comprised 2,923 proteins measured in 54,219 individuals.

To minimize potential bias from horizontal pleiotropy, cis-pQTLs were defined relative to the transcription start site (TSS). In Fenland, cis-pQTLs were defined as variants within ±1 Mb of the TSS^57^. Approximate conditional analyses were conducted to identify independent signals based on distance-based clumping, yielding 2,900 genome-wide significant cis-pQTLs across 1,557 genes (*P* < 1×10⁻¹¹). In ARIC, cis-regions were defined as ±500 kb of the TSS^59^. We retained significant independent cis-pQTLs as reported by the study, restricted to a 5% false discovery rate (FDR), resulting in 2,004 and 1,618 unique protein instruments in European American and African American datasets, respectively. Similarly, deCODE used a ±1 Mb window around the TSS^58^. We selected significant (*P* < 1×10⁻⁷) independent cis-pQTLs identified through conditional analyses. After harmonization and removal of duplicate variants by chromosomal position, 5,662 unique cis-pQTLs across 1,663 protein-coding genes (corresponding to 1,674 proteins and 1,703 SOMAmer IDs) were retained. For the UK Biobank Pharma Proteomics Project (UKB-PPP), plasma proteomic profiles were generated using the Olink Explore 3072 platform in 54,219 participants^60^. This platform includes 2,941 protein analytes targeting 2,923 unique proteins and is based on proximity extension assay (PEA) technology, in which paired antibodies labeled with complementary oligonucleotides enable detection via next-generation sequencing^60^. Instruments were restricted to cis-variants within ±1 Mb of the encoding gene, and unconditional effect estimates were used. A total of 2,057 independent cis-pQTLs meeting a Bonferroni-corrected threshold (*P* < 1.7×10⁻¹¹) were included. To ensure consistency across datasets and avoid bias introduced by conditioning, we used unconditional effect estimates for all instruments in downstream analyses.

### Expression quantitative trait loci (eQTL)

Cis-eQTL instruments were derived from summary statistics from the eQTLGen Consortium (whole blood; *n* = 31,684)^61^ and the Genotype-Tissue Expression (GTEx) Project v8^62^. In GTEx, cis-eQTLs were defined as variants located within ±1 Mb of a gene’s transcription start site (TSS)^62^. We included 48 GTEx tissues, as whole blood data were obtained from eQTLGen. Independent cis-eQTLs per gene in GTEx were identified using stepwise conditional analysis (up to five rounds) within ±1 Mb of the TSS, performed using individual-level data and adjusting for the peak variant if the association passed *P* < 1×10⁻⁴. The primary signal was defined as the unconditional association. To identify independent signals, we considered primary and conditional associations that passed *P* < 5×10⁻⁸ and extracted effect size and standard error estimates from the unconditional association. For eQTLGen, one cis-eQTL per gene was selected, defined as the variant with the lowest *P* value in the summary statistics.

Genes represented across multiple pQTL and/or eQTL sources were retained. When instrumental variants were absent from the outcome GWAS, proxy variants were identified using LD (r² ≥ 0.8) based on the 1000 Genomes Project Phase 3 European reference panel. Palindromic variants and non–protein-coding genes were excluded.

### Mendelian randomization

We evaluated the effects of cis-eQTLs and cis-pQTLs on HF, HFrEF, and HFpEF using two-sample Mendelian randomization implemented in the TwoSampleMR package (v0.5.6)^63^. For genes with ≥2 instrumental variables (IVs), MR estimates were derived using the inverse-variance–weighted (IVW) method; for genes with a single IV, the Wald ratio was applied. Analyses were performed separately for each QTL dataset (Fenland, deCODE, ARIC European, ARIC African American, UK Biobank Olink, eQTLGen, and each of the 48 GTEx tissues). Statistical significance was defined using a Bonferroni-corrected threshold based on the sum of unique gene-level tests across all QTL sources. The resulting significance thresholds were *P* < 1.57 × 10⁻⁶ for HF (n=31,822 tests), *P* < 1.57 × 10⁻⁶ for HFpEF (n=31,763 tests), and *P* < 1.57 × 10⁻⁶ for HFrEF (n=31,790 tests). For genes identified in multiple QTL datasets, effect estimates were required to be directionally concordant across QTL sources to be considered robust and advanced for follow-up analyses. Because the Bonferroni threshold is conservative, we additionally identified suggestive findings across all three HF phenotypes using a Benjamini-Hochberg false discovery rate (FDR) of 5%.

### Instrument strength and horizontal pleiotropy

To assess weak instrument bias, we calculated the proportion of variance explained (*R*^2^) and the F-statistic from the first-stage regression of the exposure on the genetic instrument. R² was derived as a function of the effect size estimate, minor allele frequency, standard error, and sample size. The F-statistic was calculated as 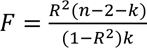, where *k* denotes the number of instruments and *n* the sample size; instruments with F < 10 were considered weak. To evaluate the consistency of instrumental variable estimates, we performed heterogeneity tests and reported corresponding p-values, with *P* < 0.05 indicating evidence of heterogeneity. Directional pleiotropy was assessed using the MR-Egger intercept test for genes with ≥3 instruments, with *P* < 0.05 indicating evidence of directional pleiotropy.

### Novelty classification

To assess the novelty of our findings, results were classified into one of three categories: novel (not previously reported for any HF phenotype), reassigned (previously reported for a different HF phenotype), or not novel (previously reported for the same phenotype). This classification was applied independently to each of the six analyses (GWAS and MR analyses for HF, HFpEF, and HFrEF), evaluated against a curated database of 12 previously published GWAS (genome-wide significance, *P* < 5 × 10⁻⁸) and MR studies (Bonferroni significance) for HF, HFrEF, and HFpEF^3–7,38,64–69^.

For GWAS findings, evidence of prior reporting was drawn from two complementary sources: a genomic proximity search identifying any previously reported variant within ±250 kb of each new locus, and gene-level annotations (cS2G, nearest gene, MAGMA, and V2G scores) matched against the same prior literature. For MR findings, novelty was assessed by matching gene names directly against those in the prior literature.

In both GWAS and MR analyses, classification followed the same phenotype-specific logic. A finding was classified as “not novel” if prior evidence existed for the same HF phenotype (e.g. a new HFpEF locus with a prior HFpEF hit in the region). It was classified as “reassigned” if prior evidence existed only for a different HF subtype (e.g. a new HFpEF locus with only prior HF or HFrEF evidence), suggesting the signal may reflect subtype-specific biology not previously identified. A finding was classified as “novel” if no prior evidence existed for any HF phenotype.

Gene assignments from each prior study were extracted as follows. For GWAS studies, nearest gene assignments were used from Henry et al. 2025 (Supplementary Table 3), Joseph et al., Lee et al. (Supplementary Table 1), Levin et al. (Supplementary Table 2), Rasooly et al. 2023, Shah et al., Wu et al. (Table 1), and Zhou et al. (Table S7); for Enzan et al. (Supplementary Table 2), both nearest gene and prioritized gene assignments were used. For MR studies, Bonferroni-significant results were used from Henry et al. 2022, Rasooly et al. 2023, Rasooly et al. 2025, Schmidt et al., Shah et al., and Zhou et al. Where a source contributed both GWAS and MR findings (Rasooly et al. 2023, Shah et al., and Zhou et al.), both were included.

### Cardiomyopathy gene sets

We further evaluated GWAS and MR gene findings against GWAS and comprehensive panel genes for hypertrophic cardiomyopathy (HCM) and dilated cardiomyopathy (DCM). For HCM GWAS genes, we used the nearest gene assignments reported in Supplementary Table 2 of a recent study^70^. For DCM, we used the nearest gene annotations reported in Supplementary Table S8 of a recent study^71^, maintaining consistency with our approach in HCM. We additionally evaluated all 30 genes from the Invitae Hypertrophic Cardiomyopathy Panel: *ACADVL, ACTC1, ACTN2, AGL, ALPK3, BAG3, CACNA1C, CPT2, CSRP3, DES, ELAC2, FHL1, FLNC, GAA, GLA, LAMP2, MTO1, MYBPC3, MYH7, MYL2, MYL3, PLN, PRKAG2, TCAP, TNNC1, TNNI3, TNNT2, TPM1, TTR*, and *VCL*. We also evaluated all 54 genes from the Invitae Dilated Cardiomyopathy and Left Ventricular Noncompaction Panel: *ABCC9, ACADVL, ACTC1, ACTN2, ALMS1, ALPK3, BAG3, CPT2, CRYAB, CSRP3, DES, DMD, DNAJC19, DOLK, DSC2, DSG2, DSP, EMD, EYA4, FKRP, FKTN, FLNC, HCN4, JUP, LAMP2, LMNA, MYBPC3, MYH7, MYLK3, PCCA, PCCB, PKP2, PLN, PPCS, RAF1, RBM20, RYR2, SCN5A, SDHA, SGCD, SLC22A5, TAZ, TBX20, TCAP, TMEM43, TMEM70, TNNC1, TNNI3, TNNI3K, TNNT2, TPM1, TTN, TTR,* and *VCL*.

### Functional enrichment analysis

Functional enrichment analysis was performed separately for HF, HFrEF, and HFpEF, incorporating all MR and GWAS results for each HF phenotype, using g:Profiler (version 0.2.4)^72^. Genes were tested for overrepresentation in Gene Ontology (GO) terms, KEGG and Reactome pathways, and other curated functional gene sets. Multiple testing was controlled using the g:Profiler set counts and sizes (SCS) correction, with adjusted *P* < 0.05 considered significant.

### Linkage disequilibrium

LD was assessed using the 1000 Genomes Project European reference panel by calculating r² between proposed variants and nearby variants. High LD was defined as r² > 0.8 and moderate LD as 0.4 ≤ r² ≤ 0.8. Analyses were performed both within our current findings– comparing variants identified for HF, HFrEF, and HFpEF– and between these variants and those reported in 12 prior genomic studies of HF, HFrEF, and HFpEF.

### ClinGen/OMIM and knockout mouse models

To assess whether our findings have been previously reported at both the phenotype and gene levels, we examined multiple orthogonal sources of evidence, including disease annotations from ClinGen and Online Mendelian Inheritance in Man (OMIM), as well as animal models (knockout and transgenic). Disease annotation data were retrieved from ClinGen and reviewed and classified for cardiovascular relevance by a cardiologist. We queried OMIM, a comprehensive catalog of human genes and genetic disorders, to identify any reported disorders or traits associated with variation in each gene. We used knockout mouse models from Mouse Genome Informatics (MGI) and the International Mouse Phenotyping Consortium (IMPC), where we extracted phenotypic data for all mutations or alleles of each gene, which were then evaluated by a cardiologist for HF and cardiomyopathy-related phenotypes.

All entries from these orthogonal sources (ClinGen, OMIM, MGI, and IMPC) were independently reviewed by two cardiologists and classified as: (1) cardiovascular phenotype, or (2) HF and cardiomyopathy phenotype. These classifications were incorporated into a per-gene summary score representing the total number of supporting sources, with positive evidence from ClinGen/OMIM and knockout mouse models contributing to the overall score.

### Evidence from secondary genes

We identified genes (referred to as “secondary genes”) directly linked to our MR and GWAS findings (or “primary genes”). These were defined using a framework introduced by MacNamara^73^, which integrates multiple interaction resources to construct a comprehensive protein–protein interaction (PPI) network, including the Human Reference Interactome Mapping Project (Lit-BM) and ligand–receptor interactions from MetaBase. To harmonize data across resources, gene identifiers were mapped to standardized gene symbols, and interactions were aggregated for each gene across all datasets. Because genes and drug targets exert downstream effects through biological networks, we systematically annotated each secondary gene using the same framework applied to primary genes. Specifically, we retrieved OMIM and ClinGen evidence, which was reviewed and classified for cardiovascular relevance by a cardiologist, as well as animal model evidence (knockout and transgenic) from MGI and IMPC, which were reviewed and classified for HF, cardiomyopathy, and cardiovascular relevance by two cardiologists. We further assessed whether secondary genes had prior evidence from HF, HFrEF, or HFpEF GWAS or MR studies, were included in HCM or DCM gene panels, or were identified in the current study. For each primary gene, we quantified the number of HF/cardiomyopathy-positive sources across all of its connected secondary genes.

### MitoCarta 3.0 Human evidence

We obtained mitochondrial evidence and functional annotations, including evidence status, sub-mitochondrial localization, and curated mitochondrial pathway assignments, for each gene using Human MitoCarta3.0^74^. Quantitative proteomics metrics were also obtained, including the number of tissues with mass spectrometry support and proteome coverage, as well as heart-specific protein abundance. In addition, subcellular localization data from the Human Protein Atlas were incorporated, including reliability scores. These annotations were used to characterize mitochondrial localization, function, and tissue-specific expression of genes implicated in HF, HFrEF, and HFpEF.

### Cardiac structure and function

For each genome-wide significant HF, HFrEF, and HFpEF variant, we evaluated associations with 25 cardiac magnetic resonance (CMR) traits to determine whether variants were also associated with cardiac structure or function at genome-wide significance (*P* < 5 × 10⁻⁸). The traits included diastolic global longitudinal strain rate (long-axis), ratio of mitral inflow early diastolic velocity (E) to early diastolic strain rate (eSR), left atrial maximum volume, LV global circumferential strain (short-axis), LV global longitudinal strain (long-axis), LV global radial strain (short-axis), LV end-diastolic volume (and indexed to body surface area), LV end-systolic volume (and indexed to body surface area), LV ejection fraction, LV filling pressure, LV mass (and indexed to body surface area), LV stroke volume (and indexed to body surface area), median global myocardial T1 relaxation time, RV global circumferential strain (short-axis), RV global longitudinal strain (long-axis), RV global radial strain (short-axis), RV end-diastolic volume (and indexed to body surface area), RV end-systolic volume, and RV stroke volume (and indexed to body surface area)^75^.

Separately, for genes prioritized by MR for HF, HFrEF, and HFpEF, we performed MR analyses using each of the 25 CMR traits as outcomes. Significance was defined using the same Bonferroni-corrected threshold applied in the primary MR discovery analysis (*P* < 1.57 × 10⁻⁶), which is more stringent than a Bonferroni-corrected threshold adjusting for the number of tests performed in this analysis. For each gene, we quantified the number of CMR traits with significant MR associations. MR beta concordance between HF, HFrEF, and HFpEF and cardiac MRI traits was determined based on whether effect directions aligned with the expected relationship of each trait with HF risk. Beta SD categories were defined within each trait by calculating the mean and standard deviation of the absolute beta coefficients (|β|), and classifying values according to their standardized distance from the trait-specific mean, using thresholds of <1 SD, 1–2 SD, and ≥2 SD.

### Body mass index

For each genome-wide significant variant, we evaluated associations with body mass index (BMI)^76^ to determine whether variants were also associated with BMI at genome-wide significance (*P* < 5 × 10⁻⁸). Separately, for genes prioritized by MR, we performed MR analyses using BMI as the outcome. Significance was defined using the same Bonferroni-corrected threshold applied in the primary MR discovery analysis (*P* < 1.6 × 10⁻⁶).

### Evidence from orthogonal sources

We aggregated evidence from multiple orthogonal sources into a composite score, including: OMIM/ClinGen annotations for HF, cardiomyopathy, or other cardiovascular-related conditions; knockout mouse model evidence for HF, cardiomyopathy, or other cardiovascular-related phenotypes; evidence from cardiac MRI traits capturing cardiac structure and function; and evidence from published GWAS/MR studies of HF and its subtypes (to evaluate replicability), as well as cardiomyopathies (dilated cardiomyopathy or hypertrophic cardiomyopathy GWAS, or a comprehensive cardiomyopathy gene panel). In addition, we performed network-based analyses to identify secondary genes connected to each primary gene. For each secondary gene, we assessed evidence of association with HF or cardiomyopathy using OMIM/ClinGen annotations and knockout mouse model data.

### Tissue-specific expression profiling

Candidate genes prioritized from GWAS and MR loci were evaluated for tissue-specific expression using RNA sequencing data from Expression Atlas^77^. Genes were categorized into five tissue groups based on physiological relevance to HF: cardiac (heart, heart left ventricle, atrial appendage), vascular (coronary artery, tibial artery), kidney (kidney, cortex of kidney), skeletal muscle, and adipose tissue. Normalized transcripts per million (TPM) values were used to quantify gene-level RNA expression across tissues; transcripts with a binned expression value of −1 (indicating below detectable threshold) were excluded. For each gene, mean and maximum TPM values were calculated across the constituent tissues within each category. Genes were ranked by RNA expression within each tissue category and classified into top 25%, middle 50%, or bottom 25% tiers relative to the genome-wide distribution of all genes with available expression data in ExpressionAtlas for that tissue category (cardiac: n = 24,025; vascular: n = 23,948; kidney: n = 24,271; skeletal muscle: n = 23,513; adipose: n = 24,367).

### Druggability assessment

Genes were annotated for therapeutic tractability using Open Targets (version 25.12)^78^, following methods previously described by our group^4^. For genes classified as druggable, corresponding compounds were retrieved from ChEMBL (v34)^79^, including approved and investigational drugs, indications, clinical development phase and mechanism of action (MoA). Translational candidates were defined as genes that are the sole known target of a compound and for which the direction of the MR effect on HF or its subtypes was concordant with the drug’s MoA (e.g., genetically predicted increased expression consistent with pharmacologic agonism). FDA-reported cardiac adverse events associated with each compound were extracted to inform safety considerations.

### Data Availability

The GWAS summary statistics generated and analyzed in this study will be available through dbGaP under study accession phs001672. The only restriction is that use of the data is limited to health/medical/biomedical purposes and does not include the study of population origins or ancestry.

### Code Availability

We used publicly available software for the analyses, and all software used is listed and described in the Methods section. Statistical analyses were conducted in R version 4.2.1. Mendelian randomization analyses were conducted using the TwoSampleMR package in R (version 0.6.6); standardization of GWAS summary statistics was conducted using MungeSumstats (release 3.20). Imputation was conducted using Minimac4 (version 1.0.0). The GWAS findings were functionally annotated using FUMA (version 1.3.8). Meta-analysis of GWAS summary statistics was conducted using METAL (version released 6 October 2020).

## Supporting information

MVP Core Acknowledgements

## Acknowledgements

We are grateful to all the MVP investigators; a list of MVP investigators can be found in the Supplementary Materials. This research is based on data from the Million Veteran Program, Office of Research and Development, Veterans Health Administration, and was supported by awards #MVP037 [BLR&D Merit Award BX005831], #MVP001 [I01-BX004821], #MVP065 [PI: Joseph], #MVP000, and Veterans Affairs Grant I01CX001922 (PI: Joseph). We also acknowledge the VA Merit Grant I01-CX001025 (PI: Wilson/Cho). This publication does not represent the views of the Department of Veterans Affairs or the United States Government. NA and HN acknowledge funding support from the Medical Research Council (MR/X020924/1).

