## Supplementary Figures for "Global genomics in over 4 million individuals prioritizes therapeutic targets for heart failure and its subtypes"

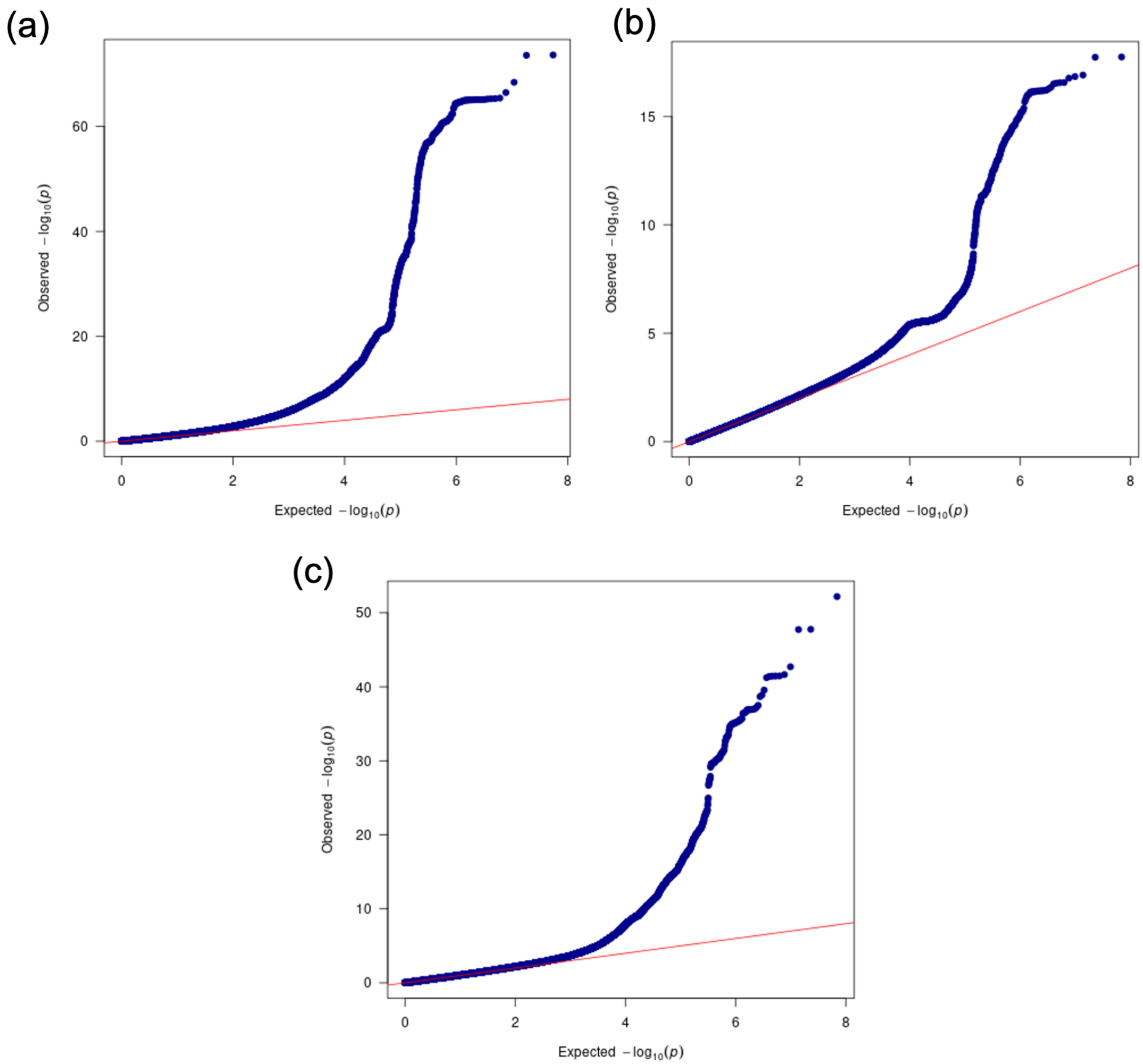


**Figure S1. Quantile-quantile (Q-Q) plots for multi-ancestry GWAS for (a) HF, (b) HFpEF, and (c) HFrEF.** Q-Q plots demonstrate the distribution of observed genome-wide association p-values to the expected distribution under the null hypothesis. Each point represents the ordered -log10(p-value) from the GWAS results plotted against the expected -log10(p-value), with the diagonal line representing null expectation. Inflation of test statistics, assessed using the genomic inflation factor ($\lambda_{GC}$), was 1.22 for HF, 1.05 for HFpEF, and 1.08 for HFrEF.

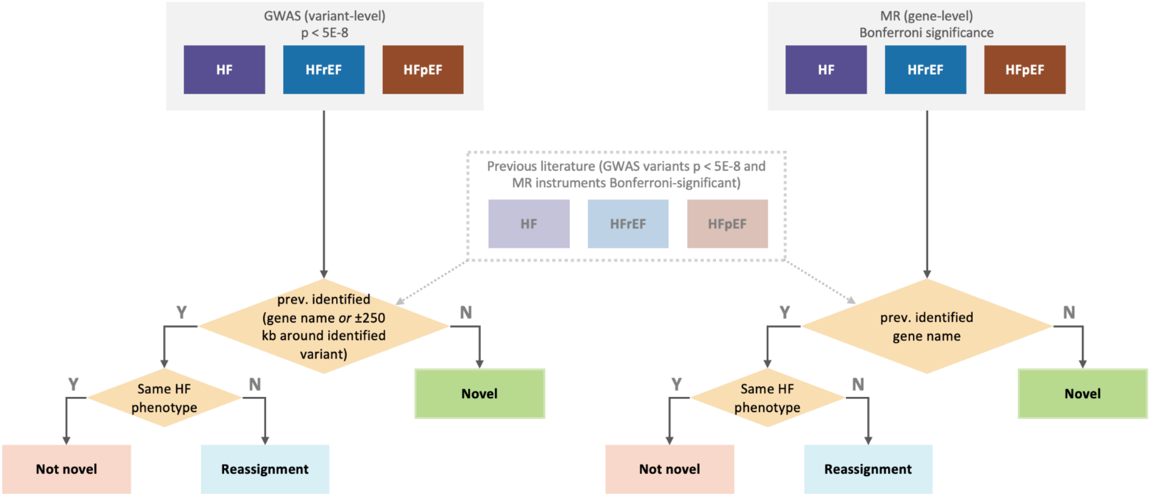


| | Not novel if prior is | Reassignment if prior is |
|---|---|---|
| HF (GWAS/MR) | HF | HFpEF or HFrEF |
| HFpEF (GWAS/MR) | HFpEF | HF or HFrEF |
| HFrEF (GWAS/MR) | HFrEF | HF or HFpEF |

**Figure S2. Novelty classification framework for identified GWAS variants and MR genes.** Findings from multi-ancestry GWAS and MR analyses of HF, HFrEF, and HFpEF were systematically classified for novelty against a curated database of 12 previously published GWAS ($P < 5 \times 10^{-8}$) and MR studies (Bonferroni significance) for HF, HFrEF, and HFpEF. For the novelty classification of the GWAS loci, evidence was drawn from two sources: gene-level annotations (cS2G, nearest gene, MAGMA, and V2G) and a genomic proximity search for any previously published HF GWAS or MR variant within ±250 kb of each identified variant. For the novelty classification of the MR gene findings, the MR gene name was matched directly against gene names reported in the published HF GWAS and MR literature. In both cases, a finding was classified as "Not Novel" if prior evidence existed for the same HF phenotype, "Reassignment" if prior evidence existed only for a different HF phenotype (e.g. a gene previously implicated in HFpEF was identified in HF, a gene previously implicated in HF was identified in HFpEF, or a gene previously implicated in HFrEF was identified in HFpEF), and "Novel" if no prior evidence was found.

(a)

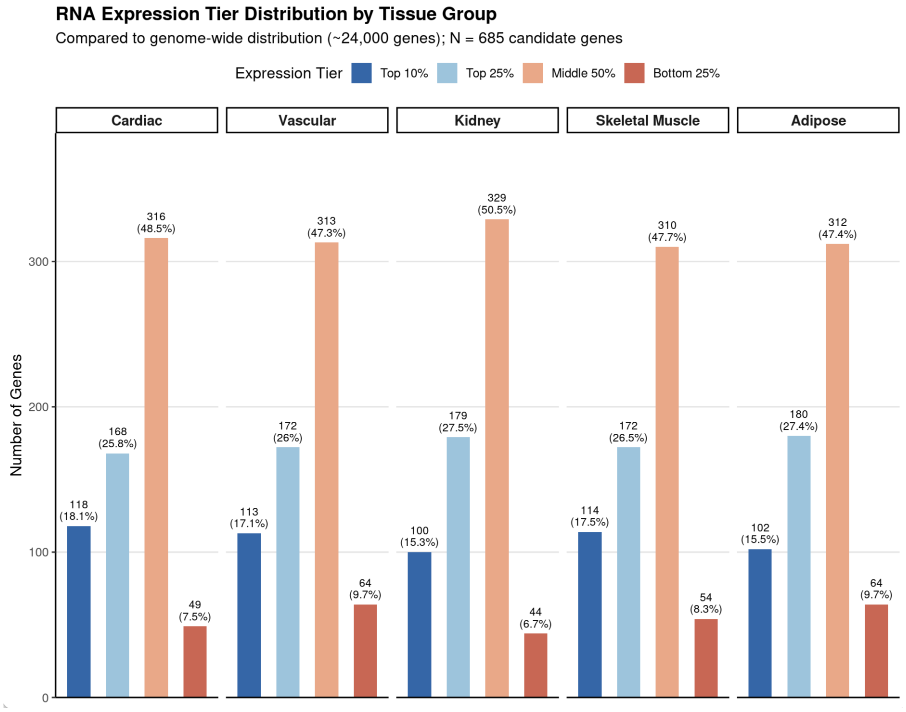
RNA Expression Tier Distribution by Tissue Group
Compared to genome-wide distribution (~24,000 genes); N = 685 candidate genes
Expression Tier
Top 10%
Top 25%
Middle 50%
Bottom 25%
Cardiac
Vascular
Kidney
Skeletal Muscle
Adipose
Number of Genes
0
100
200
300
118 (18.1%)
168 (25.8%)
316 (48.5%)
49 (7.5%)
113 (17.1%)
172 (26%)
313 (47.3%)
64 (9.7%)
100 (15.3%)
179 (27.5%)
329 (50.5%)
44 (6.7%)
114 (17.5%)
172 (26.5%)
310 (47.7%)
54 (8.3%)
102 (15.5%)
180 (27.4%)
312 (47.4%)
64 (9.7%)

(b)

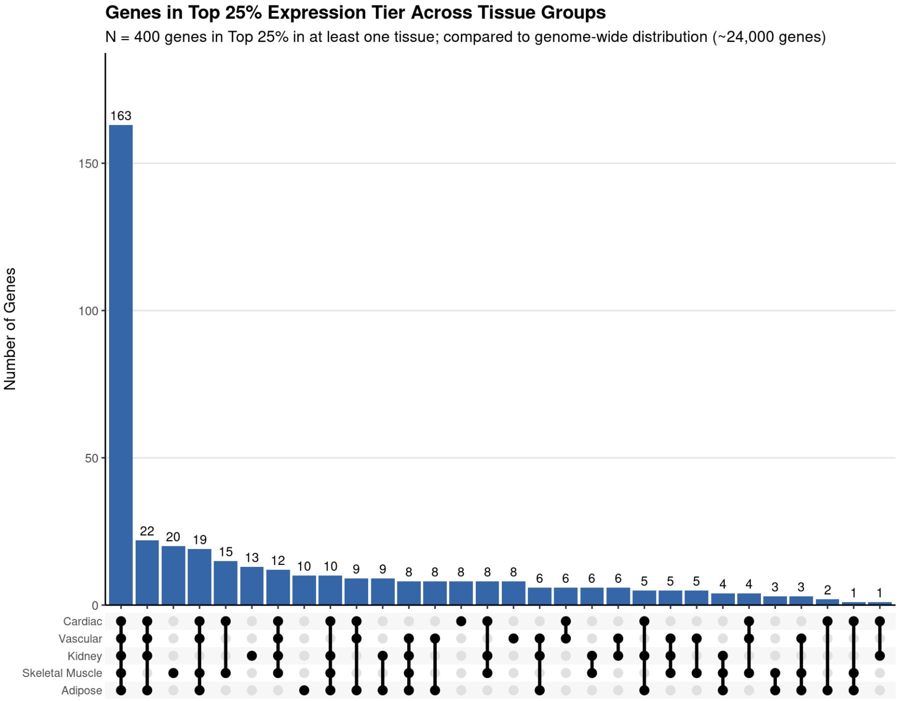
Genes in Top 25% Expression Tier Across Tissue Groups
N = 400 genes in Top 25% in at least one tissue; compared to genome-wide distribution (~24,000 genes)
Number of Genes
0
50
100
150
163
22
20
19
15
13
12
10
10
9
9
8
8
8
8
8
6
6
6
6
5
5
5
4
4
3
3
2
1
1
Cardiac
Vascular
Kidney
Skeletal Muscle
Adipose

(c)

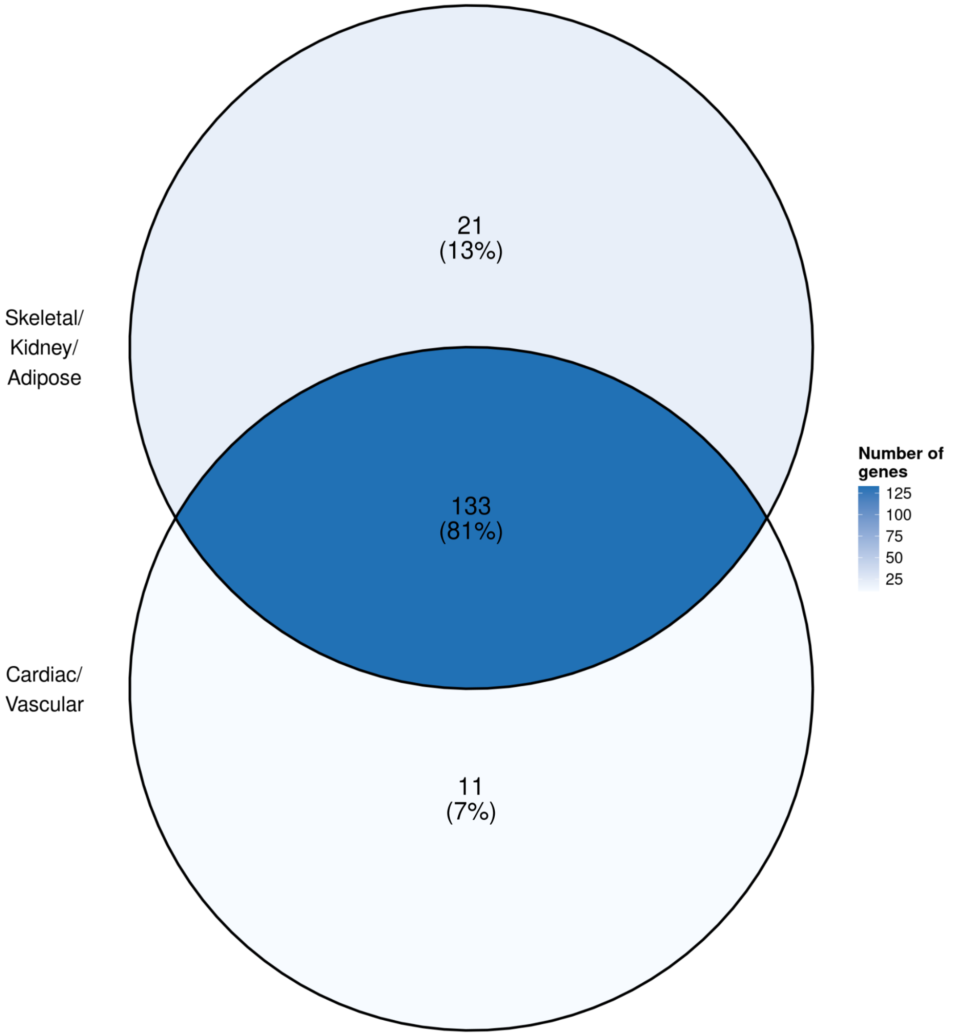
21
(13%)
Skeletal/
Kidney/
Adipose
133
(81%)
Cardiac/
Vascular
11
(7%)
Number of
genes
125
100
75
50
25

(d)

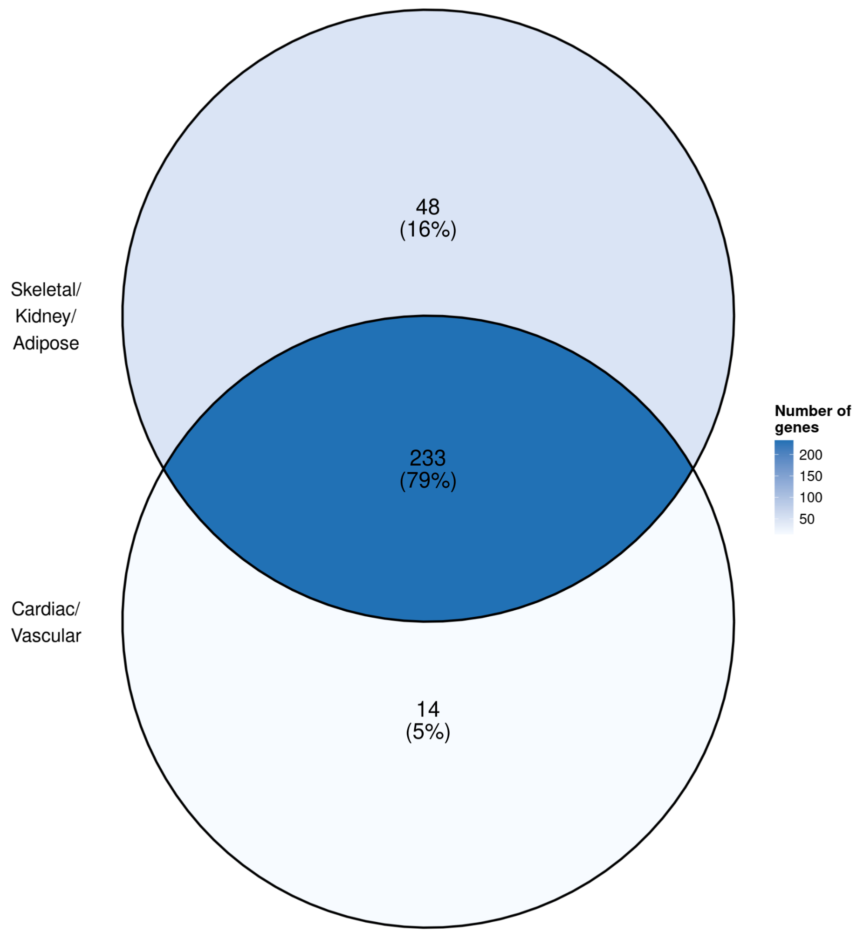


**Figure S3. Tissue-specific RNA expression enrichment of HF genes.** (a) Distribution of RNA expression tiers across major HF-related tissues for 685 candidate genes prioritized through GWAS and Mendelian randomization analyses. Gene expression was evaluated in cardiac, vascular, kidney, skeletal muscle, and adipose tissues and compared with the genome-wide distribution of approximately 24,000 protein-coding genes. Bars represent the number of candidate genes within each tissue classified into expression tiers based on tissue-specific expression rank: top 10%, top 25%, middle 50%, and bottom 25% of expressed genes. (b) UpSet plot showing overlap of candidate genes within the top 25% expression tier across cardiac, vascular, kidney, skeletal muscle, and adipose tissues. (c) Venn diagram showing the overlap between GWAS genes that fall exclusively in the top expression quartile (top 25%) in cardiac or vascular tissue ("Cardiac/Vascular") and those exclusively in the top expression quartile in skeletal muscle, kidney, or adipose tissue ("Skeletal/Kidney/Adipose"). Expression tier ("highly expressed") was defined relative to the genome-wide expression distribution across all genes assessed. (d) Same as (c), but for MR genes.

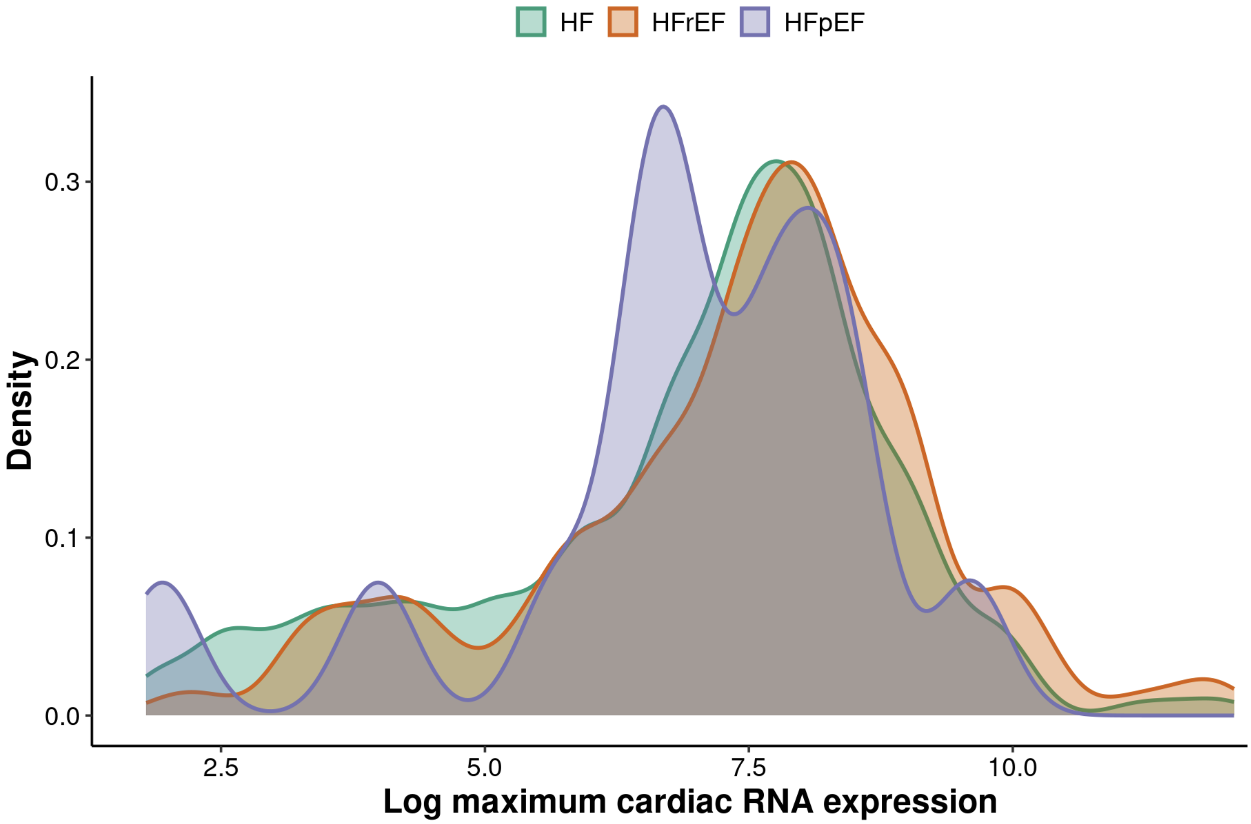


**Figure S4. Distribution of maximum cardiac RNA expression among HF, HFpEF, and HFrEF genes.** Overlaid density plots illustrate the distribution of maximum expression across the three HF phenotypes. The log-transformed maximum cardiac RNA expression (in transcripts per million, TPM) was calculated across three cardiac tissues: heart, heart left ventricle, and atrium auricular region using bulk RNA-seq data. TPM count is normalized transcript abundance for all transcripts of a given gene in each tissue.

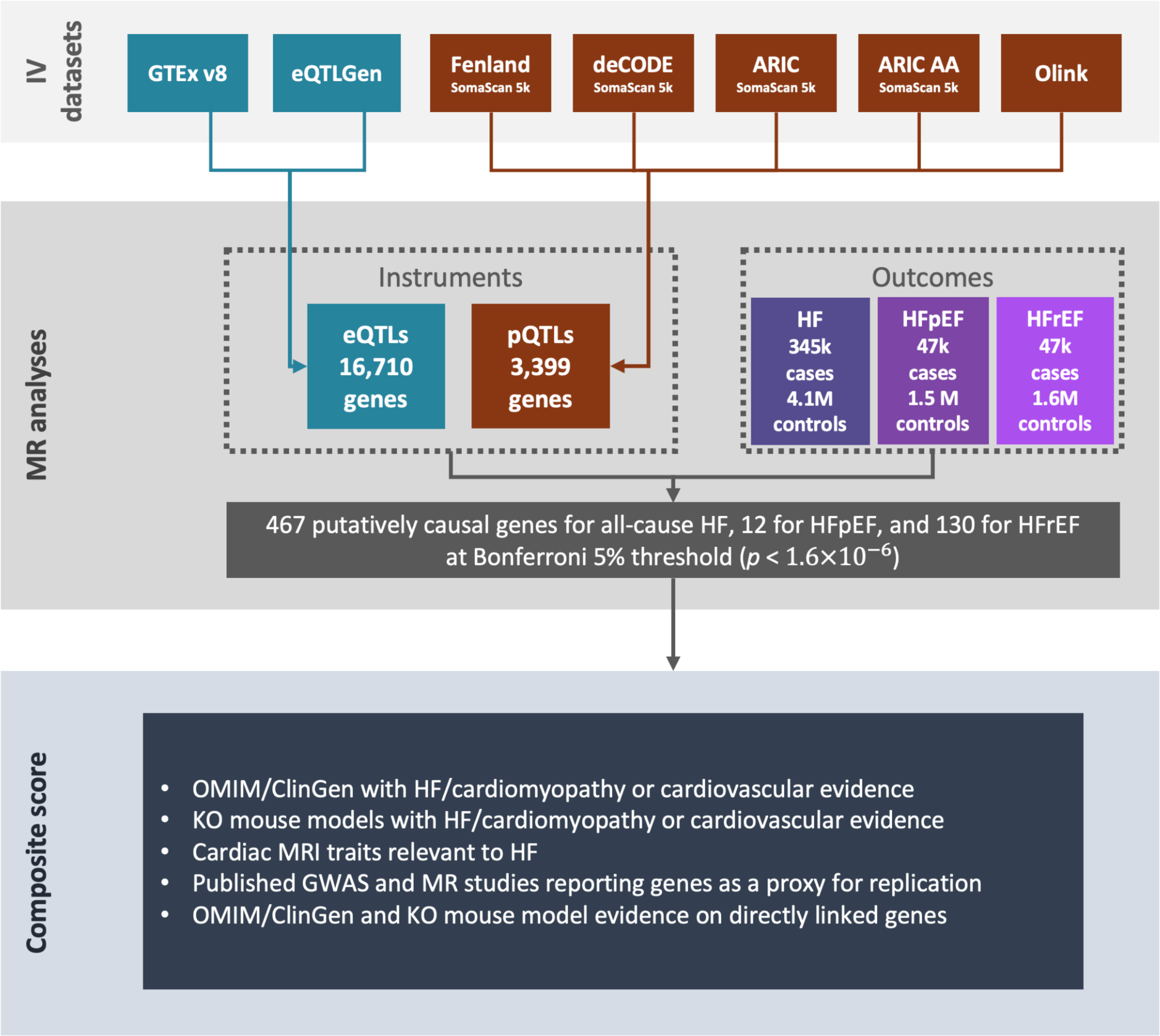


**Figure S5. Study overview for the therapeutic target profiling of HF, HFrEF, and HFpEF.** We quantified the MR effect of expression quantitative trait loci (eQTLs) from GTEx and eQTLGen and protein quantitative trait loci (pQTLs) from the Fenland, deCODE, ARIC European, ARIC African-American, and UK Biobank Pharma Proteomics Project (UKB-PPP) Olink Explore 3072 studies on all-cause heart failure (HF), HF with preserved ejection fraction (HFpEF), and HF with reduced ejection fraction (HFrEF). Findings are reported using a Bonferroni-adjusted threshold of 5% ($p$ < 1.6E-6).

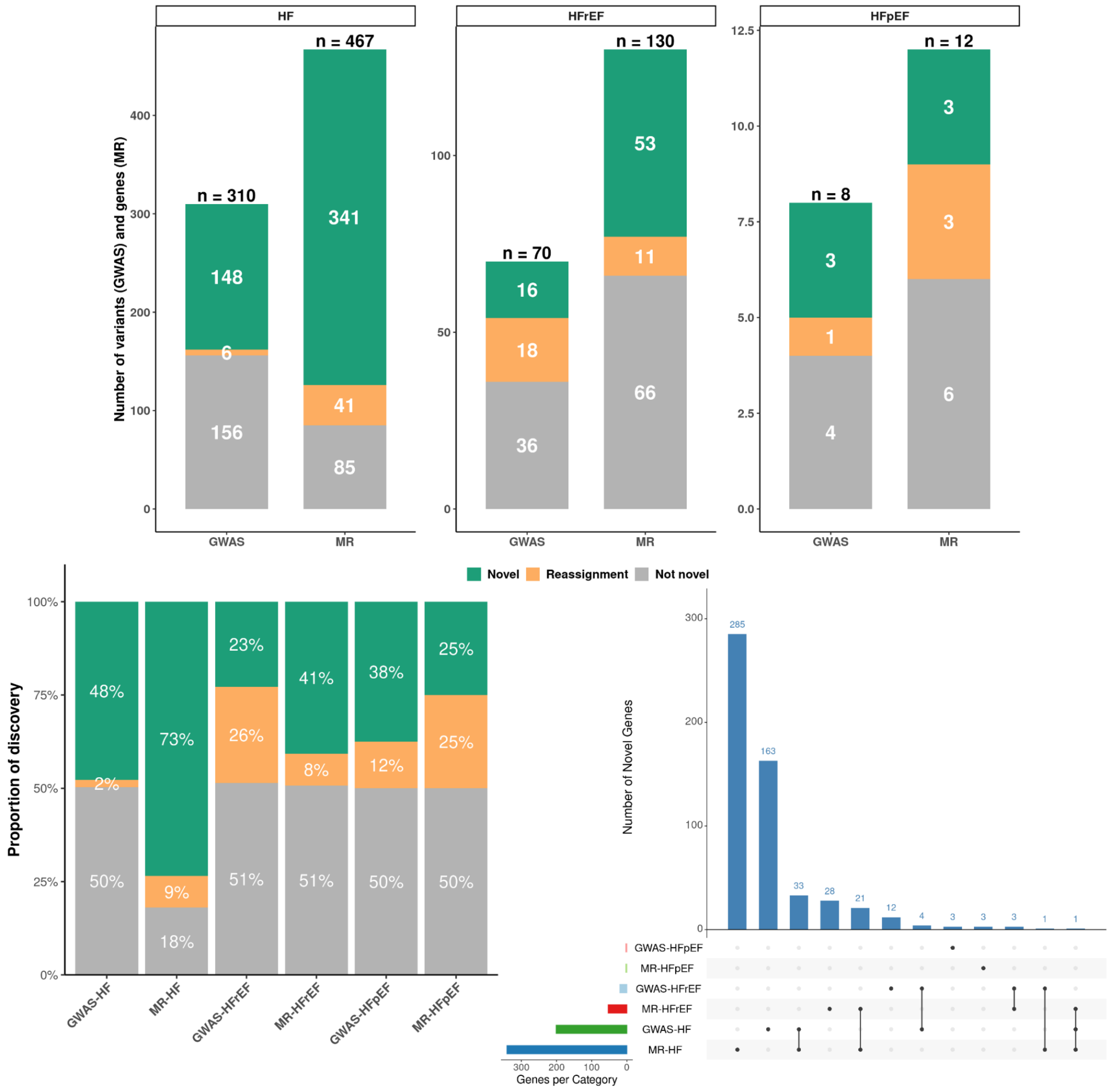


**Figure S6. Distribution and overlap of novel HF genes across phenotype and discovery method.** (a) Number of variants (GWAS) and genes (MR) for each of the six analyses. Total gene counts shown above each bar. (b) Proportion of discoveries in each of the six analyses according to novelty classification. Novelty definitions and color schemes are consistent with panel (a), and percentages are displayed within each bar. (c) UpSet plot illustrating intersections among novel genes only across the six analyses. Raw intersection counts are shown above the bars, highlighting shared and phenotype- or method-specific novel genes. For GWAS results, we used the most frequently assigned gene as the representative gene name. When a GWAS variant had multiple equally frequent gene assignments, each gene was evaluated independently when comparing overlap with MR-identified genes in this figure.

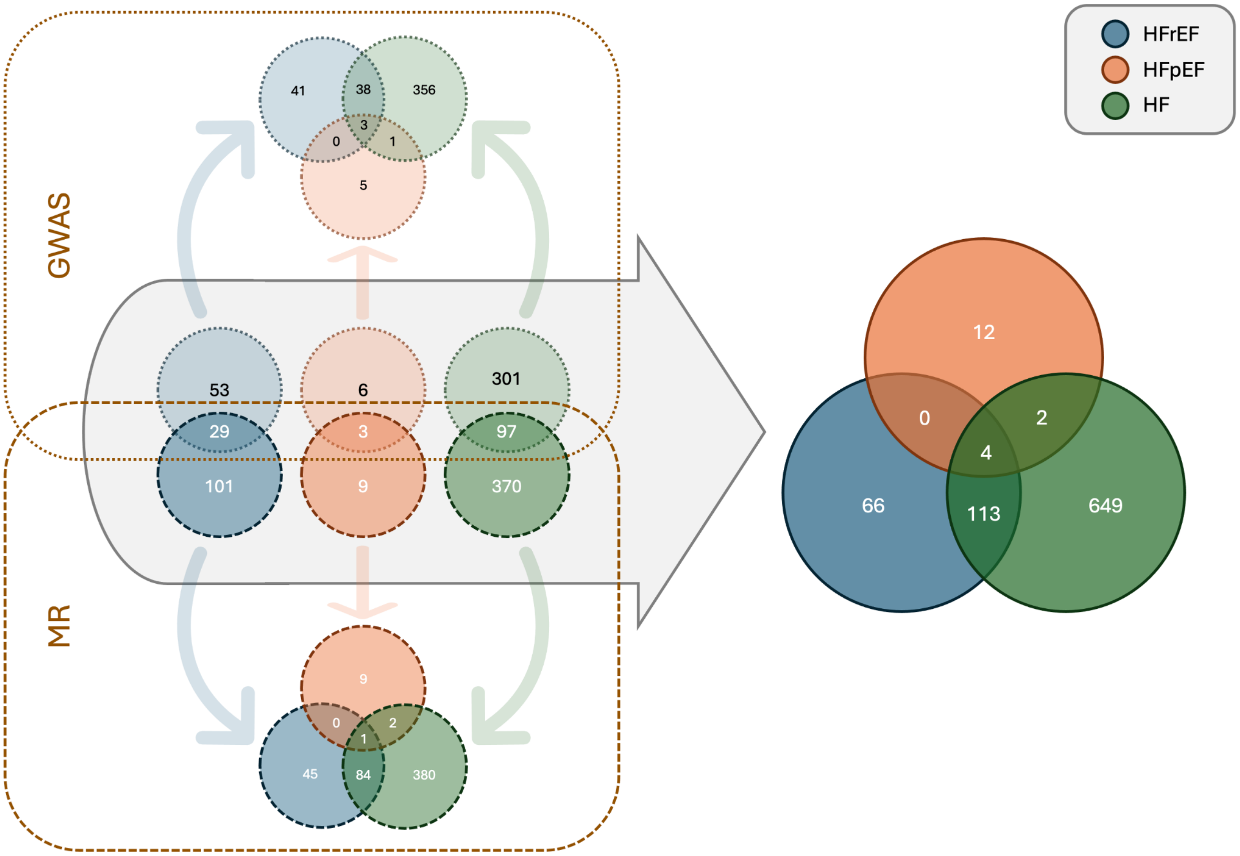


**Figure S7. Overlap of GWAS and MR genes across HF, HFrEF, and HFpEF.** Venn diagrams illustrate the overlap of candidate genes identified through GWAS and MR analyses across HF, HFrEF, and HFpEF. (Top left) Overlap of most frequently assigned GWAS genes across HF, HFrEF, and HFpEF. (Center left) Phenotype-specific overlap between discovery methods, comparing most frequently assigned GWAS genes and MR genes within HF, HFrEF, and HFpEF. (Bottom left) Overlap of MR genes among HF, HFrEF, and HFpEF. (Right) Overlap of combined most frequently assigned GWAS gene and MR-derived genes across HF, HFpEF, and HFrEF. For GWAS loci with multiple most frequently assigned gene names, all gene names were parsed individually and each unique gene name was counted once per set. In all panels, circles are color-coded by phenotype (HFpEF, orange; HF, green; HFrEF, blue).

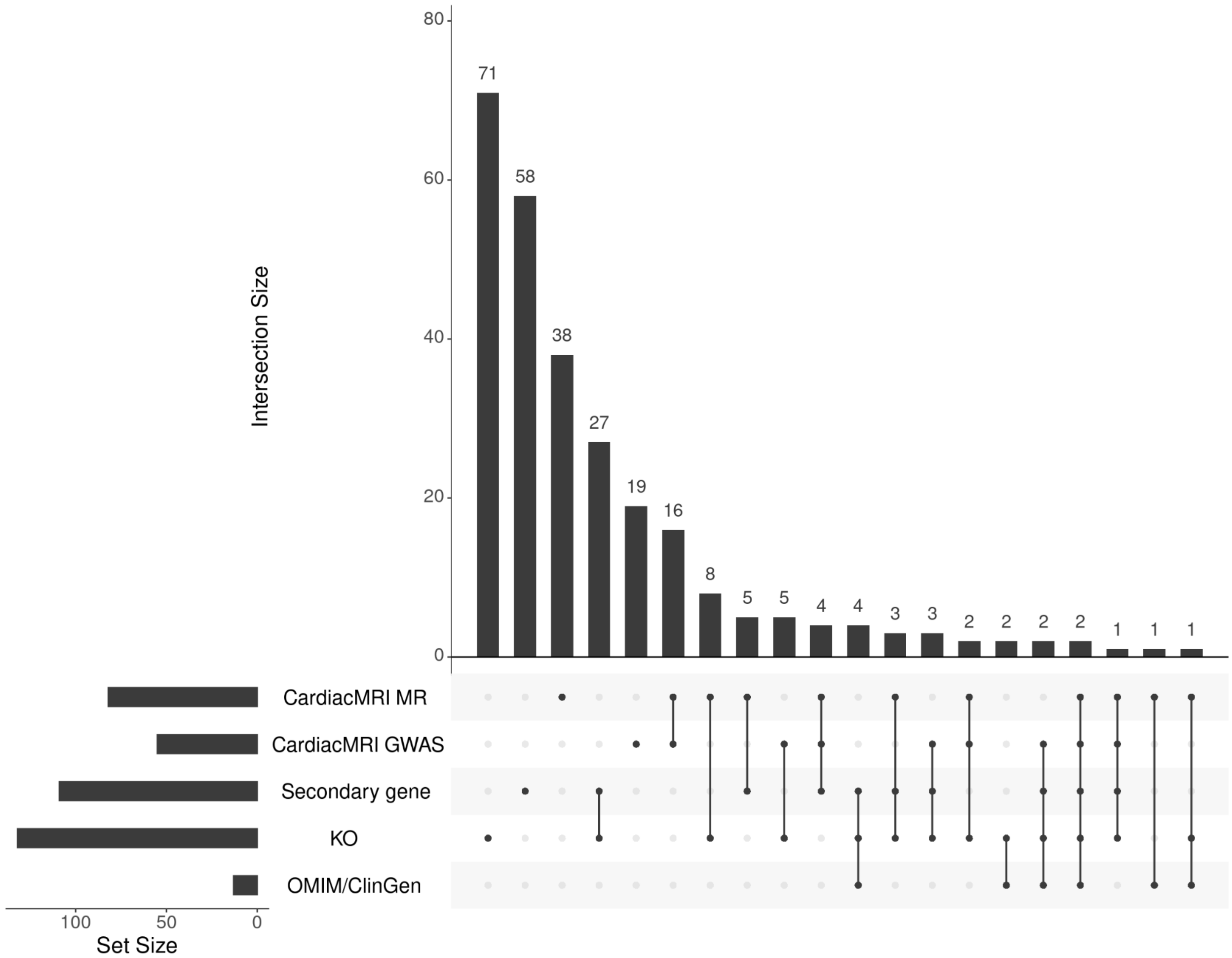


**Figure S8. Upset plot of orthogonal sources of support for GWAS and MR genes associated with HF, HFpEF, HFrEF.** The plot illustrates intersections across orthogonal lines of evidence, including cardiac MRI MR associations, cardiac MRI GWAS associations, secondary gene annotations with HF/cardiomyopathy or cardiovascular relevance, knockout mouse models with HF/cardiomyopathy or cardiovascular phenotypes, and OMIM/ClinGen cardiovascular evidence, for the most frequently assigned GWAS genes and MR-identified genes. Raw intersection counts are shown above each bar.

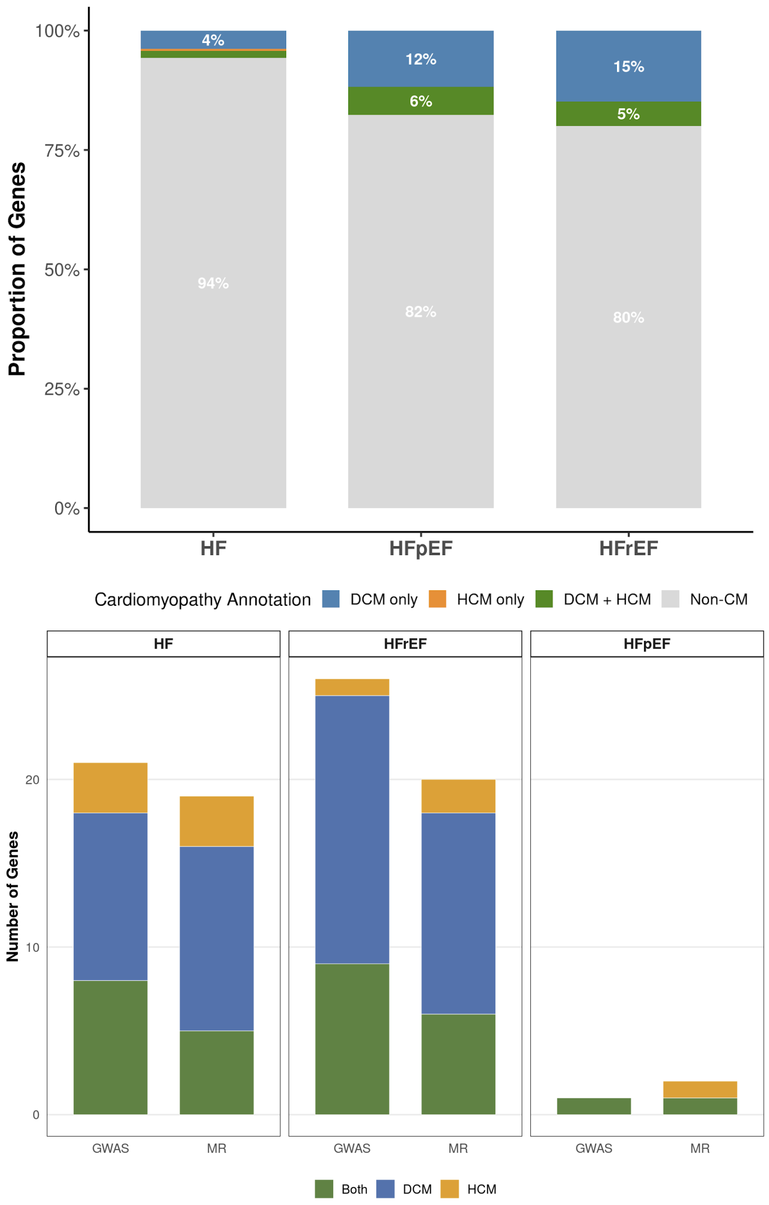


**Figure S9. Distribution of cardiomyopathy annotations among HF-associated genes.** (Top figure) Stacked bar plots show the proportion of genes associated with dilated cardiomyopathy (DCM only), hypertrophic cardiomyopathy (HCM only), both DCM and HCM (DCM + HCM), or not associated with cardiomyopathy (Non-CM) for HF, HFrEF, and HFpEF. The percentage of genes in each category is indicated within the bars. Genes from both GWAS and MR analyses are combined. (Bottom figure) Bar plot demonstrating the count of the number of genes for HF, HFpEF, and HFrEF by GWAS and MR which are associated with dilated cardiomyopathy (DCM only), hypertrophic cardiomyopathy (HCM only), both DCM and HCM (DCM + HCM). Note the DCM and HCM annotations include GWAS and comprehensive panels.
